# Better individual-level risk models can improve the targeting and life-saving potential of early-mortality interventions

**DOI:** 10.1101/2021.07.20.21260818

**Authors:** Chad Hazlett, Antonio P. Ramos, Stephen Smith

## Abstract

Infant mortality remains high and uneven in much of sub-Saharan Africa. Even low-cost, highly effective therapies can only save lives in proportion to how successfully they can be targeted to those children who, absent the treatment, would have died. This places great value on maximizing the accuracy of any targeting or means-testing algorithm. Yet, the interventions that countries deploy in hopes of reducing mortality are often targeted based on simple models of wealth or income or a few additional variables. Examining 22 countries in sub-Saharan Africa, we illustrate the use of flexible (machine learning) risk models employing up to 25 generally available pre-birth variables from the Demographic and Health Surveys. Using these models, we construct risk scores such that the 10 percent of the population at highest risk account for 15-30 percent of infant mortality, depending on the country. Successful targeting in these models turned on several variables other than wealth, while models that employ only wealth data perform little or no better than chance. Consequently, employing such data and models to predict high-risk births in the countries studied flexibly could substantially improve the targeting and thus the life-saving potential of existing interventions.

## Introduction

Goal 3.2 of the Sustainable Development Goals (SDG) seeks to “end preventable deaths of newborns and children under five years of age, with all countries aiming to reduce neonatal mortality to at least as low as 12 per 1,000 live births and under-5 mortality to at least as low as 25 per 1,000 live births.” Despite large overall improvements, including a 44% reduction in child mortality globally from 2000 to 2015, progress has varied widely both between and within countries,^1,2,3^ with under-5 mortality exceeding 80 per 1,000 live births in some countries of sub-Saharan Africa. Further, improvements in national averages can mask large and widening gaps between sub-populations, with more privileged groups improving their health status faster than others.^4,5^

Many of these deaths are preventable as they can be avoided with low-cost interventions such as oral rehydration therapy, malaria prevention and treatment, vaccines (e.g., against measles), and nutritional support.^6,7^ However, a critical feature of any life-saving intervention is that it can only save the lives of those who would have otherwise died. Even a “miracle drug” that can counteract any cause of death can only reduce mortality when given to children who would have died without it. Because of this, interventions that cannot be given universally must be carefully targeted to those at the highest risk of mortality (absent the intervention) to have an effect. While this point may seem obvious, in the context of statistically rare events, it has enormous implications because it can severely limit any intervention’s theoretical maximum life-saving potential. Consider a country with an infant mortality rate of 80 deaths per 1,000 births. Normatively, this is far too high, yet this rate of 0.8% means that intervention given to a random set of individuals can save at most 0.8% of those to whom it is given.

Worse, in practice, such an intervention will only save the lives of a fraction of those who would have otherwise died. We call this fraction the *efficacy rate* (it is sometimes known as the “probability of necessity and sufficiency” in the causal inference literature). Suppose the efficacy rate is 10%, meaning the intervention would save the lives of one-tenth of those who would have otherwise died. An intervention given at random could save the lives of only 0.8% * 10% = 0.08% of those given to whom it is given. By contrast, if such an intervention *with the same efficacy rate* could instead be targeted to a group with a higher risk of mortality, say 10%, then it would save the lives of 10% * 10% = 1% of recipients, saving 12.5 times as many lives.^1^ In this way, the lives saved by a hypothetical intervention with a given efficacy rate are limited by and proportional to the mortality risk in the group to which it is given. Efforts to improve targeting should thus be treated with the same urgency as efforts to improve the efficacy of the interventions. Accordingly, our paper seeks to demonstrate the use of tools for better estimating which births are at the highest risk of early mortality.

Unfortunately, the importance of targeting has not been well recognized by recent and current public health practices in many cases. Although early mortality can have many causes and can be related to many risk factors, countries facing high early mortality often target related interventions based on one or a few factors, including income, wealth, or poverty, and employing simple statistical models or checklists. Similarly, academic research into models for early mortality that look beyond income, poverty, or wealth remains rare and not designed to demonstrate how practical, national-level systems could target interventions to high-risk births. A recent, partial exception finds, first, that poverty is a poor proxy for the risk of early mortality in Brazil and India, much as we find below for countries in sub-Saharan Africa.^8^ Further, for births in India, they find that using just four risk factors in a linear model roughly doubles the fraction of infant deaths that can be identified in the highest risk quintile compared to models relying on a proxy for poverty.

We demonstrate the potential for tools that better estimate which births are at the highest risk of early mortality, using flexibly estimated, individual-level risk scores based only on a range of commonly available pre-birth data, which can improve the targeting of life-saving interventions against infant mortality. Specifically, we estimate mortality risk in individual infants (under one year of age) in 22 countries in sub-Saharan Africa, employing now-standard machine learning models to identify predictive relationships that may have eluded more rigid regression models. This flexibility is especially valuable as we provide these models access to a wider range of risk factors than in previous work. We also employ only risk factors that can generally be known or anticipated before a child’s birth, as practical targeting systems may be limited to such data. As is standard in many machine learning applications, we employ cross-validation and held-out test sets to protect against overly optimistic performance claims due to over-fitting. However, rather than employing the mathematically complicated, non-intuitive performance metrics commonly employed in the forecasting literature, we rely on a simpler set of metrics designed to be easily interpreted by policymakers and to directly speak to the potential impact of a hypothetical policy intervention.

## Materials and Methods

### Data Sources

We employ data from the Demographic and Health Surveys (DHS), as augmented and prepared for analysis by the Integrated Public Use Microdata Series (IPUMS), henceforth referred to as the IPUMS DHS data. Each survey we used was the most recent one available via IPUMS, when our study began. Like other work studying early mortality using the IPUMS DHS data (e.g.,^9,10^), we use the “child module” which contains data on births for children born up to five years before the interview of their mother. Though interviews are conducted with mothers, one observation in the data represents each child’s birth. This module restricts our attention to children under five, but contains more predictive variables than the “all births” module. More importantly, it would be inadvisable to use data collected on the mother, household, malaria prevalence, location, etc., in the year of the DHS data collection to predict early mortality from birth many years before.

Originally, the IPUMS DHS data contained surveys on 28 sub-Saharan African countries. We consider geographic information to be critical for these models since (i) even coarse geographic indicators can be highly predictive of risk and reduce the need for other, harder-to-measure variables associated with location, (ii) we considered malaria prevalence a potentially important variable, but because it is computed for a 10km area around the DHS sample cluster centroid, it is only available in surveys that collected geographic coordinates, and (iii) the ability to target interventions partly by geography is convenient for policymakers. Unfortunately, in six countries (Burundi, Chad, Ethiopia, Liberia, Namibia, and Sudan), the IPUMS DHS data did not include geographic indicators even at the district level. This limits us at present to surveys from 22 countries: Angola (2015), Benin (2011), Burkina Faso (2010), Cameroon 2011, Democratic Republic of Congo (2013), Cote d’Ivoire (2011), Ghana (2014), Guinea (2012), Kenya (2014), Lesotho (2014), Madagascar (2008), Malawi (2016), Mali (2012), Mozambique (2011), Niger (2012), Nigeria (2013), Rwanda (2014), Senegal (2017), Tanzania (2015), Uganda (2016), Zambia (2013), Zimbabwe (2015). Table 1 provides additional information on the available data within each country.

### Non-geographic variables

Our outcome is infant mortality, indicated by death before reaching one year of age. For predictors or risk factors (we use the terms interchangeably) we limit ourselves to predictors that (i) would be reasonably available to policymakers and health workers in-country, and (ii) are measurable prior to the birth of the child, rather than depending on the health or other features of the child once borne. The latter constraint is in place so that these risk factors could be used to calculate risk in advance and deploy interventions to households, clinics, or regions with high risk.

The resulting risk factors we consider are: sibling death at an age less than 1 yr., sibling death older than 1 year, mother education, birth month (sin and cosine), sex (female), a rural indicator, local malaria prevalence, maternal age, previous pregnancy terminated, clean water access, birth order (log), mother’s age at first union, number of bednets (log(+1)), floor type, age of head of household, female household head, minority religion, christian, or muslim, unimproved toilet, pit toilet, clean cooking fuel, and within-country wealth percentile (and its square). The source data from which these are constructed are described in Table 3 of the Supplement.

Note that the IPUMS DHS data do not indicate how an individual or household is classified relative to the poverty line in a given country, nor does it attempt to measure income as such. Thus, rather than including any locally defined poverty measure, we employ the wealth-based indices provided by DHS. This measure is an index for within-country wealth, imputed by DHS using principal components analysis over several variables, including ownership of radios, televisions, and other domestic equipment; electricity and clean water; type of materials used in the walls, floor, and roof; and the type of toilet in the household. This score is calculated at the household level. The only requirement we make of this measure is that, within a given country, it provides an imperfect but potentially informative measure of relative wealth. Since this index is constructed within each country, and we train each country’s risk model separately, there is no expectation or need for cross-country comparability.

### Modeling

We estimate infant mortality risk for each birth in the data set. In each country, we separately run two sets of models. One is a “wealth model”, described below, which reveals what can be learned from the available wealth data alone. The other set contains flexible machine-learning approaches that use the predictors described above to estimate individual-level mortality risk. These include random forest (rf), extreme gradient boosting over trees (xgb), kernel regularized least squares (krls), and elastic-net regularized logistic regression (elastic-logit).

Both rf^11^ and xgb^12^ are tree-based approaches. The simplest tree-based approaches are classification and regression trees (cart^13^), which recursively partition observations into groups based on one cutoff value of the predictor variables so that each group has similar values of the outcome. While such single-tree models are easy to interpret, they often perform poorly, and small input data changes can dramatically affect the model and results. The rf and xgb methods improve on single-tree approaches by combining many trees. For rf, this is achieved by effectively averaging the estimates of many trees, each employing only a random subset of the input variables and trained on a random subset of the observations. As applied here, each forest contains 500 trees, and the number of variables randomly sampled for each tree split (mtry) was set for each country’s model using cross-validation. Under xgb, this is similarly achieved by combining many trees, but by beginning with one tree, adding the predictions of another tree trained on the errors of the prior, and then repeating this process hundreds or thousands of times. We use cross-validation for each country to determine (i) the learning rate (*η ∈* (0, 1), where smaller values help to prevent overfitting), (ii) the maximum depth of the trees (max depth), (iii) the proportion of the variables to be considered for tree construction (colsample bytree), (iv) the proportion of observations from the training set used for modeling (subsample), and (v) the number of iterations for the boosting procedure (nrounds). The remaining parameters were set at common default values: the minimum requirement of prediction improvement before selecting a more complex model (*γ*) was set to 0, and min child weight, where higher values restrict the depth of each tree based on a measure of the homogeneity of the labels within the nodes (a sign of overfitting), was set to 1.

**Table 1:** Data availability by country.

| Country | N original | N complete | N deaths | Geography | Unavailable |
| --- | --- | --- | --- | --- | --- |
| Angola | 11024 | 10829 | 472 | province (18) | age first marriage, malaria |
| Benin | 10490 | 10199 | 471 | department (12) |  |
| Burkina Faso | 11728 | 10905 | 752 | region (13) |  |
| Cameroon | 9041 | 8450 | 564 | region or city (12) |  |
| Congo (D.R.) | 14384 | 12731 | 785 | province (11) | previous child death(s) |
| Cote d'Ivoire | 5945 | 5211 | 421 | none |  |
| Ghana | 4548 | 4203 | 189 | none |  |
| Guinea | 5422 | 5223 | 396 | region (8) | cooking fuel |
| Kenya | 16557 | 15110 | 589 | none | previous pregnancy ended |
| Lesotho | 2360 | 2127 | 145 | district (10) | bednets, malaria |
| Madagascar | 9657 | 9229 | 459 | region (22) | cooking fuel |
| Malawi | 13633 | 13231 | 563 | region (3) | malaria |
| Mali | 8043 | 7913 | 475 | region (6) | cooking fuel, malaria |
| Mozambique | 8493 | 8026 | 537 | province (11) |  |
| Niger | 9650 | 9612 | 586 | region (8) | religion, malaria |
| Nigeria | 24222 | 23559 | 1686 | geopolitical zone (6) |  |
| Rwanda | 6055 | 5350 | 168 | province (5) | cooking fuel |
| Senegal | 9585 | 9371 | 401 | region (14) | malaria |
| Tanzania | 8007 | 7688 | 319 | district (10) | cooking fuel, religion, malaria |
| Uganda | 12089 | 11619 | 484 | district (112) | cooking fuel, malaria |
| Zambia | 10646 | 9811 | 483 | none |  |
| Zimbabwe | 4852 | 4630 | 236 | province (10) | malaria |

*Note:* Summary of data by country. The DHS data initially include **N original** observations in each country, which falls to **N complete** when we remove missing values on the included variables. **Geography** indicates what type of geographic information, if any, is available and the number of units in parentheses. All other variables described in text are used in each country except where otherwise indicated under **Unavailable**.

Kernel-based regularized least squares (krls^14^) is a regression and classification technique that employs kernels. Such approaches effectively build a model by leveraging information about similar observations. Each observation is treated as a potential “fence post” in the data, and the model learns how being closer to or further from each such location in the data influences the expected probability of mortality. We employ a Gaussian kernel as the measure of similarity using default values.^14^

Elastic-net logistic regression (elastic-logit^15^) is similar to conventional logistic regression. While ordinary logistic regression chooses coefficients *β_j_* on each variable *j* to maximize the likelihood by minimizing the negative log-likelihood, elastic-net logistic regression minimizes the negative log-likelihood *plus* a penalty term that helps to regularize or “shrink” the model, preventing overfitting. In our case that penalty term is equal to 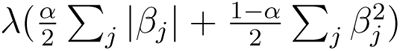. The parameters *α* and *λ* are chosen by cross-validation. This constitutes a compromise between what is known as ridge regression (which is penalized by the *l*_2_ north, 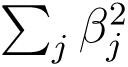), and LASSO (penalized by the *ℓ*_1_ norm, 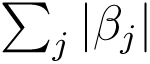). By including the *ℓ*_1_ norm in the penalty, elasticnet can set some coefficients to exactly 0, that is, selecting some variables while dropping others.

Finally, we construct a simple ensemble model in which each predicted probability is the unweighted average of the probabilities produced by elastic-logit, rf, xgb, and krls. This has two primary benefits. First, such ensembles often perform as well as the best sub-models and sometimes better. Second, it allows us to commit to reporting results for a single model—the ensemble model—as our premier estimate of the predictive power of these models on the test data. This is preferable to reporting the best-performing model on the test set, which can be misleading because such a choice effectively means “training on the test set”.

### Wealth model

To determine how much information is available in the wealth data alone and to provide a baseline performance level for comparison, we also model mortality using only wealth data. To allow a flexible model for wealth, we regress mortality on indicators for wealth, equivalent to finding the mean mortality level in each wealth decile. In evaluating model performance, we can consider how being in the highest-risk decile, quartile, or other groups is related to mortality risk. There is no presumption that risk is necessarily decreasing or even monotonic in wealth.

We note that this is a wealth model, not a poverty model, based on poverty assessments made by countries, usually based on income. Neither income nor poverty assessments themselves are available in the DHS data. This model therefore is not a direct proxy for poverty-line-based means-testing sometimes employed. That said, our wealth model may reasonably provide an optimistic indication of what can be predicted by income or poverty-based approaches. Household income is time-sensitive but is likely to generate risks of early mortality when it remains low on average over a long period, in which case wealth will also drop. Moreover, financial shocks such as large expenses or losses may have little impact on income but substantially on the quality of life, access to healthcare, and poverty as locally understood (see e.g.^16^). Our wealth model is also more flexible and less coarse than poverty-based targeting systems, which typically assign individuals through a binary system (e.g. above or below the poverty line). Again this is intended to be a favorable choice for the wealth model so that we can later consider its demonstrated performance likely to be over-rather than under-stated.

### Sample splitting and cross-validation

Before fitting any model, we split the data into training and test sets. For each country, 80% of observations are placed in a training and validation set, while 20% are retained in a held-out testing set. For the models described above, we tune hyperparameters using either 10-fold cross-validation within the training set (for elastic-logit, rf, xgb) or leave-one-out cross-validation (for krls). The cross-validation procedure also gives a preliminary assessment of model performance. Once all modeling choices were finalized based on this cross-validation, those models were “frozen”. We completed a paper draft using these cross-validation results to ensure all decisions were finalized. Only then did we run the model on the test set to obtain an honest out-of-sample performance assessment without “training on the test set” by revisiting any decisions.

### Variable importance

Our models only seek to determine the best risk estimate possible given a set of predictors, and the answers say nothing of the causal effect of those predictors on the risk of mortality. Indeed, many risk factors, such as religion or the type of roof, are valuable not because they cause mortality but because they are expected to signal the presence of other, unobserved factors that cause higher mortality. In other words, confounding is expected and hoped for in these models.

It is valuable, nevertheless to understand which variables the models deem most important in estimating the risk to assess the model’s credibility, improve future models, or focusing public health resources on data collection. Unfortunately, for many machine learning models, it is more difficult to characterize the model’s behavior than with simpler regression approaches like OLS or logit, in which a small number of parameters fully characterizes the model. That said, readily available measures of variable importance for different models can provide useful insights. Specifically, for elastic-net, we report how often the model chose a given variable and the proportion of time their coefficient is positive. For random forest, the (scaled) “variable importance” describes what fraction of the time a given variable was chosen to be included in the classification trees over which each random forest model aggregates.

### Performance metrics

Finally, we employ simple, policy-relevant metrics rather than more esoteric and difficult-to-interpret metrics often relied upon in machine learning tasks to evaluate our approach. A model’s *recall* (also referred to as sensitivity) is the proportion of actual deaths included in the group we consider to be at risk. What we term “recall10” is defined by first determining the group of births in the top risk decile (hence the “10”) according to our model, then computing the fraction of all actual infant deaths accounted for by this group. In policy terms, this corresponds to a case in which we have resources to target 10% of the population and wish to know how much of the total mortality risk would be covered in that group. An ineffective risk assessment that randomizes risk would produce a recall10 of 0.10 (10%). By contrast, an effective system points us to a top risk decile that accounts for well over 10% of mortality. Similarly, we report recall20, which tells us the fraction of early deaths within the top risk quintile. For recall10 and recall20, we also report “efficiency gain”, which tells us how many times more effective the ensemble model is than the benchmark wealth model. We consider hypothetical scenarios where it would be difficult for health ministries to target an intervention to more than 10% or 20% of the birth population due to budget constraints and practical exigencies. Finally, note that because we are interested in how these models would perform when applied across the population of births in each country, these recall measures are constructed using sampling weights (v005).

## Results

### Performance of the wealth model

Table 2 and 3 provide the primary results, showing the proportion of mortality among the top 10% (Tables 2) or 20% (Table 3) highest predicted risk individuals in the test set. The online supplement provides the same performance metrics but using cross-validated training set results. Those results are extremely similar to the test set results reported here, indicating little over-fitting in the training process.

Focusing first on wealth as a predictor, on the one hand, the average recall10 of the wealth model is only 0.12—slightly better than chance. In 15 of 22 countries, the recall10 is no greater than .12, and in some countries, it is very low (e.g., just 0.03 in Tanzania). On the other hand, there are two countries with recall10 of 0.16 (Benin and Kenya), and two with very promising recall10 values over 0.20 (Uganda, Rwanda).

Similarly, Table 3 shows that in the countrywide average, only 24% of infant mortality is captured in the top quintile of risk according to the wealth-only model (average recall20 is 0.24). Rwanda is again an exception, with the top quintile accounting for 48% of mortality. Lesotho similarly has a recall10 of 0.34. In these two countries the wealth model out-performs the more flexible models attempted. On the other hand, the recall20 of the wealth model is no better than chance (*≤* 0.20) in Cote d’Ivoire, Guinea, Madagascar, Mali, Mozambique, Niger, Tanzania, and Zimbabwe.

**Table 2:** Recall10 results, test set.

| Survey | wealth | elastic-logit | rf | xgb | krls | ensemble | efficiency gain |
| --- | --- | --- | --- | --- | --- | --- | --- |
| Angola 2015 | 0.12 | 0.22 | 0.22 | 0.19 | 0.19 | 0.22 | 1.91 |
| Benin 2011 | 0.16 | 0.23 | 0.31 | 0.20 | 0.22 | 0.23 | 1.47 |
| Burkina Faso 2010 | 0.12 | 0.15 | 0.21 | 0.16 | 0.15 | 0.20 | 1.67 |
| Cameroon 2011 | 0.07 | 0.16 | 0.23 | 0.15 | 0.15 | 0.22 | 3.13 |
| Congo (D.R.) 2013-14 | 0.14 | 0.19 | 0.19 | 0.19 | 0.16 | 0.20 | 1.41 |
| Cote d'Ivoire 2011 | 0.12 | 0.29 | 0.18 | 0.29 | 0.27 | 0.26 | 2.20 |
| Ghana 2014 | 0.14 | 0.11 | 0.22 | 0.16 | 0.08 | 0.19 | 1.40 |
| Guinea 2012 | 0.11 | 0.14 | 0.22 | 0.15 | 0.13 | 0.19 | 1.67 |
| Kenya 2014 | 0.16 | 0.21 | 0.16 | 0.12 | 0.19 | 0.20 | 1.21 |
| Lesotho 2014 | 0.14 | 0.17 | 0.14 | 0.14 | 0.10 | 0.17 | 1.25 |
| Madagascar 2008 | 0.09 | 0.10 | 0.15 | 0.10 | 0.13 | 0.21 | 2.38 |
| Malawi 2016 | 0.10 | 0.11 | 0.20 | 0.19 | 0.14 | 0.17 | 1.73 |
| Mali 2012 | 0.07 | 0.18 | 0.21 | 0.23 | 0.22 | 0.23 | 3.14 |
| Mozambique 2011 | 0.07 | 0.07 | 0.18 | 0.11 | 0.10 | 0.15 | 2.00 |
| Niger 2012 | 0.12 | 0.15 | 0.13 | 0.15 | 0.13 | 0.15 | 1.29 |
| Nigeria 2013 | 0.12 | 0.20 | 0.21 | 0.19 | 0.24 | 0.25 | 2.15 |
| Rwanda 2014 | 0.24 | 0.21 | 0.09 | 0.21 | 0.21 | 0.21 | 0.88 |
| Senegal 2017 | 0.11 | 0.17 | 0.28 | 0.29 | 0.16 | 0.32 | 2.89 |
| Tanzania 2015 | 0.03 | 0.19 | 0.14 | 0.14 | 0.16 | 0.21 | 6.50 |
| Uganda 2016 | 0.20 | 0.14 | 0.24 | 0.23 | 0.25 | 0.25 | 1.26 |
| Zambia 2013 | 0.10 | 0.19 | 0.21 | 0.14 | 0.20 | 0.20 | 1.90 |
| Zimbabwe 2015 | 0.09 | 0.09 | 0.21 | 0.19 | 0.17 | 0.15 | 1.75 |
| Average | 0.12 | 0.17 | 0.20 | 0.18 | 0.17 | 0.21 | 2.05 |

### Beyond wealth: Gains from richer models

The elastic-net logit, rf, xgb, and krls models all augment the wealth model by adding the additional variables described above. Overall these models perform similarly to each other, and in all but a few cases, considerably better than the wealth-only model. According to these models, averaging by country, the top risk deciles account for 17-21% of mortality (recall10); the top risk quintile accounts for 30-34% of mortality (recall20). On average, the ensemble model performs better than any of the individual models capturing 21% of the mortality in its top risk decile (recall10), and 34% in the top risk quintile (recall20) when averaged across countries.

Compared to the wealth model, the ensemble model, on average, captures over twice as much of the mortality in its top risk decile (efficiency gain for recall10) and 1.5 times the mortality in its top risk quintile (efficiency gain for recall20). No single country always shows the highest recall10 across models, nor the lowest. Moreover, within given countries, the ensemble model outperforms the constituent models in many cases, does as well as the top one or two models in many others, and is never the worst-performing model in that country. Thus, relying on the ensemble model yields better results than any other model on average and safeguards against cases where individual models perform poorly in a given country.

**Table 3:** Recall20 results, test set.

| Survey | wealth | elastic-logit | rf | xgb | krls | ensemble | efficiency gain |
| --- | --- | --- | --- | --- | --- | --- | --- |
| Angola 2015 | 0.31 | 0.41 | 0.34 | 0.36 | 0.38 | 0.44 | 1.41 |
| Benin 2011 | 0.29 | 0.36 | 0.47 | 0.34 | 0.39 | 0.46 | 1.59 |
| Burkina Faso 2010 | 0.23 | 0.35 | 0.27 | 0.31 | 0.41 | 0.36 | 1.54 |
| Cameroon 2011 | 0.24 | 0.25 | 0.32 | 0.24 | 0.27 | 0.29 | 1.22 |
| Congo (D.R.) 2013-14 | 0.24 | 0.28 | 0.32 | 0.30 | 0.31 | 0.35 | 1.45 |
| Cote d'Ivoire 2011 | 0.20 | 0.48 | 0.30 | 0.48 | 0.40 | 0.48 | 2.35 |
| Ghana 2014 | 0.22 | 0.16 | 0.22 | 0.30 | 0.19 | 0.24 | 1.12 |
| Guinea 2012 | 0.18 | 0.27 | 0.38 | 0.24 | 0.34 | 0.33 | 1.86 |
| Kenya 2014 | 0.26 | 0.27 | 0.32 | 0.23 | 0.31 | 0.32 | 1.23 |
| Lesotho 2014 | 0.34 | 0.31 | 0.28 | 0.21 | 0.17 | 0.24 | 0.70 |
| Madagascar 2008 | 0.19 | 0.26 | 0.31 | 0.24 | 0.26 | 0.30 | 1.59 |
| Malawi 2016 | 0.24 | 0.21 | 0.30 | 0.28 | 0.22 | 0.28 | 1.15 |
| Mali 2012 | 0.16 | 0.35 | 0.34 | 0.34 | 0.35 | 0.39 | 2.47 |
| Mozambique 2011 | 0.19 | 0.17 | 0.30 | 0.20 | 0.25 | 0.24 | 1.30 |
| Niger 2012 | 0.19 | 0.23 | 0.20 | 0.27 | 0.24 | 0.26 | 1.41 |
| Nigeria 2013 | 0.21 | 0.39 | 0.35 | 0.37 | 0.40 | 0.40 | 1.93 |
| Rwanda 2014 | 0.48 | 0.30 | 0.33 | 0.36 | 0.36 | 0.36 | 0.75 |
| Senegal 2017 | 0.26 | 0.36 | 0.38 | 0.39 | 0.30 | 0.41 | 1.57 |
| Tanzania 2015 | 0.11 | 0.32 | 0.29 | 0.33 | 0.25 | 0.27 | 2.43 |
| Uganda 2016 | 0.26 | 0.29 | 0.33 | 0.38 | 0.35 | 0.42 | 1.60 |
| Zambia 2013 | 0.26 | 0.32 | 0.32 | 0.29 | 0.30 | 0.29 | 1.12 |
| Zimbabwe 2015 | 0.17 | 0.26 | 0.23 | 0.30 | 0.21 | 0.34 | 2.00 |
| Average | 0.24 | 0.30 | 0.31 | 0.31 | 0.30 | 0.34 | 1.54 |

### Variable importance

Here we characterize the sets of variables most important to forming the risk forecasts, as indicated by variable selection choices in the elastic-net logistic regression and the established “variable importance” measure in the random forest (Table 4). In the elastic-net logistic regression model, every variable was included in at least 23% of the country-level models. A few variables, though, are included especially often: most models included the previous death of a sibling in the first year (86%) or the death of any previous sibling at any point (76%). 77% of country-level models included the child’s gender; 73% included an urban/rural indicator. Over two-thirds (68%) of models included the mother’s years of education, and many included the birth month (73% use sine-transformed birth month; 55% use cosine-transformed birth month). Maternal age was included in 64% of models.

For random forest, the “scaled variable importance” describes what fraction of the time a given variable was chosen to be included in the classification trees over which each random forest model aggregates. Compared to elastic-net logistic regression, the random forest was somewhat more selective, with over half of the variables appearing in fewer than 15% of the regression trees. A few variables were rarely included, such as clean cooking fuel (3%) and minority religion (5%). By contrast, maternal age was included nearly every time (99.6%), followed closely by wealth percentile and its square (93%, 94%). The latter set is particularly notable, as wealth percentiles were among the *least* commonly employed in the elastic-net logit models. Other important variables included malaria incidence rate in the area (92%), age of household head (77%), and mother’s age at first marriage or union (63%).

**Table 4:** Variable Importance.

| variable | elastic<br>survival | proportion<br>positive | rf variable<br>importance |
| --- | --- | --- | --- |
| sibling death (< 1 yr.) | 0.86 | 1.00 | 0.11 |
| mother education | 0.77 | 0.06 | 0.39 |
| sin(month) | 0.73 | 0.69 | 0.46 |
| female | 0.73 | 0.00 | 0.17 |
| sibling death | 0.71 | 1.00 | 0.11 |
| rural | 0.64 | 0.71 | 0.10 |
| malaria | 0.62 | 0.75 | 0.92 |
| maternal age | 0.59 | 0.08 | 1.00 |
| prev. pregnancy ended | 0.57 | 0.75 | 0.12 |
| clean water | 0.55 | 0.58 | 0.14 |
| cos(month) | 0.55 | 0.33 | 0.48 |
| log(birth order) | 0.55 | 0.25 | 0.54 |
| mother age first union | 0.52 | 0.45 | 0.63 |
| log(num bednets) | 0.52 | 0.09 | 0.38 |
| safe floor | 0.50 | 0.18 | 0.11 |
| age household head | 0.50 | 0.64 | 0.77 |
| female household | 0.50 | 0.55 | 0.11 |
| minority religion | 0.50 | 0.70 | 0.06 |
| christian | 0.35 | 0.43 | 0.08 |
| muslim | 0.35 | 0.71 | 0.06 |
| unimproved toilet | 0.32 | 0.86 | 0.09 |
| clean cooking fuel | 0.31 | 0.20 | 0.05 |
| (wealth percentile)<sup>2</sup> | 0.27 | 0.00 | 0.94 |
| pit toilet | 0.23 | 0.60 | 0.10 |
| wealth percentile | 0.23 | 0.60 | 0.93 |

## Discussion

Across 22 countries in sub-Saharan Africa, wealth-based measures alone were fairly predictive of early mortality risk in a few countries (notably Rwanda, Uganda, and Lesotho) but weakly predictive in others and on average. On average, individuals in the highest 10% and 20% risk groups, according to these models, accounted for only 12% and 24% of infant mortality. When wealth information is combined with other risk factors in flexible models, it often aids in predicting individual-level risk of early mortality. On average across countries, the 10% of births with the highest risk, as predicted by the ensemble model, account for 21% of deaths, and the 20% at highest risk account for 34%. Compared to the wealth-only models, each country’s ensemble model, on average, would have identified about twice as many deaths in the top risk decile and 1.5 times as many in the top 20% risk group. We note that the relatively weak relationship found between mortality risk and wealth in many countries is indeed a *within*-country phenomenon: it is quite possible that taken across countries, differences in wealth would more strongly predict mortality. However, the nature of the analysis and the wealth data in the DHS (which is indexed to be valid only within-country) leave us examining how risk and wealth relate within a given country.

The various models used to incorporate these risk factors—a regularized logistic regression (elastic-logit), random forest (rf), extreme gradient boosting (xgb), and kernel regularized least squares (krls)—have similar average performance. However, some models exceed others in particular countries. Using the average of predictions from these (ensemble) generates better performance while protecting against cases where individual models performed poorly in particular countries. Particularly important to many models were variables regarding the previous death of siblings in the first year, the mother’s age and education, malaria prevalence, and the child’s gender. The value of these variables as predictors says nothing of their causal influence or importance. Rather, they are likely to be predictive not because they cause or prevent mortality but because they signal the presence or absence of other important unmeasured factors that do.

These findings are consistent with previous findings that births at high risk of death exist across all socioeconomic groups and that, for various purposes, combine multiple risk factors and show that doing so improves our ability to identify the higher-risk births.^17,8,18^ They also speak to a large body of literature in medicine and public health that develops risk scores to identify those at risk of some event (e.g.,^19,20^). To our knowledge, flexible machine learning approaches using numerous pre-birth predictors have not previously been used to develop birth-level mortality risk scores in sub-Saharan Africa or elsewhere.

### What recall10/20 represents

One reason to use recall10 or recall20 is that it lends itself to a simple interpretation of model performance, rooted in a hypothetical scenario involving an intervention with some fixed efficacy rate and fixed budget. Consider an intervention that a given country can afford to administer to only 10% of births. Ideally, this would be targeted to the 10% at the highest risk of early mortality. Let us suppose it has a fixed efficacy rate, which, as defined above, is the probability of saving a life that would have otherwise been lost. In a country with *N_births_* births per year, the number of lives saved per year would be efficacy *∗ Pr*(death*|* high risk) *∗ N_births_/*10. Applying Bayes’ rule, this is equal to recall10 *∗* efficacy*∗Pr*(death)*∗N_births_* where *Pr*(death) is the overall mortality rate expected for children born in that year (absent the intervention). Notice that this (hypothetical) number of lives saved is proportional to recall10. By contrast, measures commonly used for classification accuracy in forecasting work, such as “f-scores” or or the “area under the curve” on a receiver operator characteristic curve (AUC-ROC), are more difficult to connect to a substantive policy-oriented interpretation.

To complete this hypothetical example, using recent estimates of the number of births and baseline mortality rates in each of these countries,^21^ the number of lives saved by an intervention with an efficacy of 5% would be 841 thousand for the wealth model, but 1.61 million for the ensemble model. In simpler terms, the efficiency gain of roughly two for these models compared to the wealth model (or, roughly, random targeting)) translates into approximately twice as many lives saved by an intervention with any particular fixed efficacy. Note that the average efficiency gain (2.3) is not exactly the ratio of lives saved because the latter takes the relative birth rates and mortality in each country into account.

### Implications and implementation

The good news demonstrated here is that using flexible and inclusive modeling techniques can greatly improve our targeting ability and consequently produce multiplicative increases in the potential lives saved by a given intervention. Nevertheless, our analysis was only a demonstration in the general case, using widely available data. For a country to realize these gains, it will encounter practical exigencies that can increase and decrease the potential life-saving gains relative to our example. On the one hand, the real-world implementation of such an approach would be better and more appropriate than ours because the agency charged with constructing such a model can best choose to collect the variables that (i) are most likely to be predictive; (ii) are most feasible to collect widely; and (iii) are practical for purposes of targeting through existing administrative systems. On the other hand, such systems may not be prepared to collect some of the variables used here at high frequency. Further, the nature of an intervention and logistical limitations ultimately restrict the degree to which an intervention can be targeted. Nevertheless, targeting any intervention at any level can benefit from improved individual-level risk estimation since these individual-level risks can be aggregated to form the best predictions at any level. Concretely, at one extreme, suppose village health workers distributing a given intervention are positioned to enter specific data about individuals and determine their eligibility for an intervention. This would allow fully individualized risk-based targeting. Similarly, health workers could be instructed to use instead a heuristic scoring system (e.g. a checklist), which could be designed to approximate a well-fitted individual-level risk model. On the other hand, many interventions cannot be individually targeted, and are targeted to geographic areas (e.g., the placement of health facilities, decisions about where to spray for mosquitoes or to improve sanitation, etc). In these cases, individual-level risk scores still provide the ideal basis for targeting, as they can be aggregated at will to determine “area average risks” or “area total risk” for targeting. Our results suggest that in many countries, such an approach would offer larger improvements over making such decisions using local poverty or wealth measures.

### Limitations and Opportunities

Our work has several important limitations. First by focusing on individual-level risk of early mortality we demonstrate how the life-saving potential of *any* intervention—even one that can unfailingly prevent death by every cause—is severely limited by the accuracy of the targeting system used to determine who gets it. While improving risk modeling has proven here to be an effective way to amplify life-saving potential in this sense, future research could look beyond this theoretical maximum life-saving potential, considering, for example, the need for interventions that address specific causes of death and how this limits the life-saving potential of targeting or requires targeting systems sensitive to the cause of death addressed by each of several interventions. This would require a dataset that records the cause of death, which the IPUMS DHS data does not.

Second, while we aimed to rely only on the types of predictors that are already widely available through existing IPUMS DHS data, unfortunately, a few countries did not have surveys even in recent decades (Sudan) or lacked important basic variables such as district-level or lower geographic information and malaria prevalence (Burundi, Chad, Ethiopia, Liberia, Namibia). This limits generalization of our findings to areas with the lowest data availability but also points to the need to identify or build suitable data sources for these countries.

Third, the DHS sampling procedure attempts to produce a sample representative of the population. If these are not as representative as hoped, then while the results still reflect the predictive power of these models in some population of each country, the results may not be representative of these countries as a whole.

Finally, and most broadly, the central limitation is that not all deaths can be predicted, at least with the data we considered readily available in most countries. In general, we find our models for the highest 10% risk group capture only 20-30% of mortality. This is beneficial and worth using for targeting since the effectiveness of an intervention is proportional to this recall rate, as shown above. However, it leaves much unpredicted mortality spread across the lower-risk groups. Thus, when resources can be made available, programs must still be targeted to a much wider group to capture a large fraction of births that will die. Relatedly, in the future and with appropriate data, approaches like ours could be refined if information on the cause of death were available. Associating each birth to a particular cause of death could generate models that suggest not only which births to target but also which intervention(s) to make available to them.

## Data Availability

The data is available via Demographic and Health Surveys.

https://www.dhsprogram.com/

## Availability of data and materials

The data used for this analysis can be found in the Integrated Public Use Microdata Series, available at IPUMS). Data need to be requested but are publicly available. Code to reproduce all results in this paper can be found at [repository information to be linked when accepted for publication].

---

**Table 1:** Recall10 results, cross-validation on training set.

| Survey | wealth | elastic-logit | rf | xgb | krls | ensemble | efficiency gain |
| --- | --- | --- | --- | --- | --- | --- | --- |
| Angola 2015 | 0.13 | 0.19 | 0.22 | 0.18 | 0.19 | 0.21 | 1.62 |
| Benin 2011 | 0.15 | 0.31 | 0.27 | 0.29 | 0.31 | 0.32 | 2.16 |
| Burkina Faso 2010 | 0.09 | 0.16 | 0.18 | 0.16 | 0.15 | 0.17 | 1.98 |
| Cameroon 2011 | 0.11 | 0.12 | 0.20 | 0.14 | 0.12 | 0.22 | 2.06 |
| Congo (D.R.) 2013-14 | 0.13 | 0.14 | 0.20 | 0.16 | 0.18 | 0.21 | 1.63 |
| Cote d'Ivoire 2011 | 0.11 | 0.23 | 0.25 | 0.20 | 0.23 | 0.24 | 2.13 |
| Ghana 2014 | 0.15 | 0.20 | 0.12 | 0.17 | 0.17 | 0.18 | 1.22 |
| Guinea 2012 | 0.17 | 0.15 | 0.15 | 0.15 | 0.15 | 0.17 | 1.00 |
| Kenya 2014 | 0.15 | 0.15 | 0.19 | 0.12 | 0.13 | 0.17 | 1.13 |
| Lesotho 2014 | 0.08 | 0.09 | 0.17 | 0.07 | 0.15 | 0.14 | 1.63 |
| Madagascar 2008 | 0.11 | 0.23 | 0.26 | 0.24 | 0.24 | 0.29 | 2.52 |
| Malawi 2016 | 0.10 | 0.16 | 0.20 | 0.16 | 0.18 | 0.19 | 1.91 |
| Mali 2012 | 0.10 | 0.20 | 0.23 | 0.19 | 0.22 | 0.26 | 2.55 |
| Mozambique 2011 | 0.07 | 0.11 | 0.23 | 0.15 | 0.13 | 0.21 | 3.18 |
| Niger 2012 | 0.09 | 0.13 | 0.18 | 0.16 | 0.14 | 0.20 | 2.11 |
| Nigeria 2013 | 0.11 | 0.17 | 0.18 | 0.18 | 0.17 | 0.21 | 1.87 |
| Rwanda 2014 | 0.16 | 0.19 | 0.17 | 0.10 | 0.13 | 0.15 | 0.95 |
| Senegal 2017 | 0.11 | 0.20 | 0.22 | 0.25 | 0.20 | 0.26 | 2.37 |
| Tanzania 2015 | 0.08 | 0.12 | 0.18 | 0.17 | 0.14 | 0.20 | 2.55 |
| Uganda 2016 | 0.14 | 0.16 | 0.17 | 0.17 | 0.17 | 0.16 | 1.19 |
| Zambia 2013 | 0.09 | 0.15 | 0.19 | 0.19 | 0.18 | 0.22 | 2.54 |
| Zimbabwe 2015 | 0.11 | 0.14 | 0.17 | 0.17 | 0.13 | 0.17 | 1.58 |
| Average | 0.12 | 0.17 | 0.20 | 0.17 | 0.17 | 0.21 | 1.90 |

**Table 2:** Recall20 results, cross-validation on training set.

| Survey | wealth | elastic-logit | rf | xgb | krls | ensemble | efficiency gain |
| --- | --- | --- | --- | --- | --- | --- | --- |
| Angola 2015 | 0.25 | 0.34 | 0.35 | 0.36 | 0.35 | 0.38 | 1.52 |
| Benin 2011 | 0.28 | 0.45 | 0.46 | 0.47 | 0.45 | 0.49 | 1.76 |
| Burkina Faso 2010 | 0.21 | 0.30 | 0.30 | 0.29 | 0.28 | 0.31 | 1.48 |
| Cameroon 2011 | 0.22 | 0.25 | 0.32 | 0.26 | 0.24 | 0.35 | 1.61 |
| Congo (D.R.) 2013-14 | 0.24 | 0.27 | 0.29 | 0.30 | 0.31 | 0.34 | 1.40 |
| Cote d'Ivoire 2011 | 0.20 | 0.37 | 0.33 | 0.37 | 0.36 | 0.42 | 2.04 |
| Ghana 2014 | 0.32 | 0.29 | 0.26 | 0.27 | 0.28 | 0.29 | 0.89 |
| Guinea 2012 | 0.30 | 0.26 | 0.32 | 0.28 | 0.29 | 0.32 | 1.08 |
| Kenya 2014 | 0.28 | 0.26 | 0.30 | 0.21 | 0.26 | 0.30 | 1.07 |
| Lesotho 2014 | 0.17 | 0.26 | 0.28 | 0.16 | 0.26 | 0.24 | 1.42 |
| Madagascar 2008 | 0.26 | 0.37 | 0.37 | 0.39 | 0.37 | 0.43 | 1.68 |
| Malawi 2016 | 0.19 | 0.31 | 0.30 | 0.29 | 0.30 | 0.34 | 1.84 |
| Mali 2012 | 0.20 | 0.34 | 0.36 | 0.36 | 0.36 | 0.38 | 1.88 |
| Mozambique 2011 | 0.17 | 0.22 | 0.35 | 0.30 | 0.28 | 0.34 | 1.93 |
| Niger 2012 | 0.18 | 0.30 | 0.29 | 0.26 | 0.26 | 0.30 | 1.67 |
| Nigeria 2013 | 0.24 | 0.33 | 0.32 | 0.33 | 0.33 | 0.34 | 1.43 |
| Rwanda 2014 | 0.28 | 0.36 | 0.30 | 0.24 | 0.28 | 0.32 | 1.13 |
| Senegal 2017 | 0.28 | 0.35 | 0.33 | 0.34 | 0.36 | 0.36 | 1.27 |
| Tanzania 2015 | 0.15 | 0.22 | 0.29 | 0.27 | 0.24 | 0.29 | 1.87 |
| Uganda 2016 | 0.26 | 0.28 | 0.31 | 0.30 | 0.26 | 0.30 | 1.18 |
| Zambia 2013 | 0.17 | 0.31 | 0.30 | 0.33 | 0.32 | 0.34 | 2.01 |
| Zimbabwe 2015 | 0.20 | 0.27 | 0.33 | 0.31 | 0.25 | 0.33 | 1.66 |
| Average | 0.23 | 0.30 | 0.32 | 0.30 | 0.30 | 0.34 | 1.54 |

Table 3 describes the variables used in the analyses. While the variables shown here include some that we have transformed or processed already, these constitute the raw material from which variables used in the analysis are formed (see description in Materials and Methods and in Table 4 of the main text.)

**Table 3:**
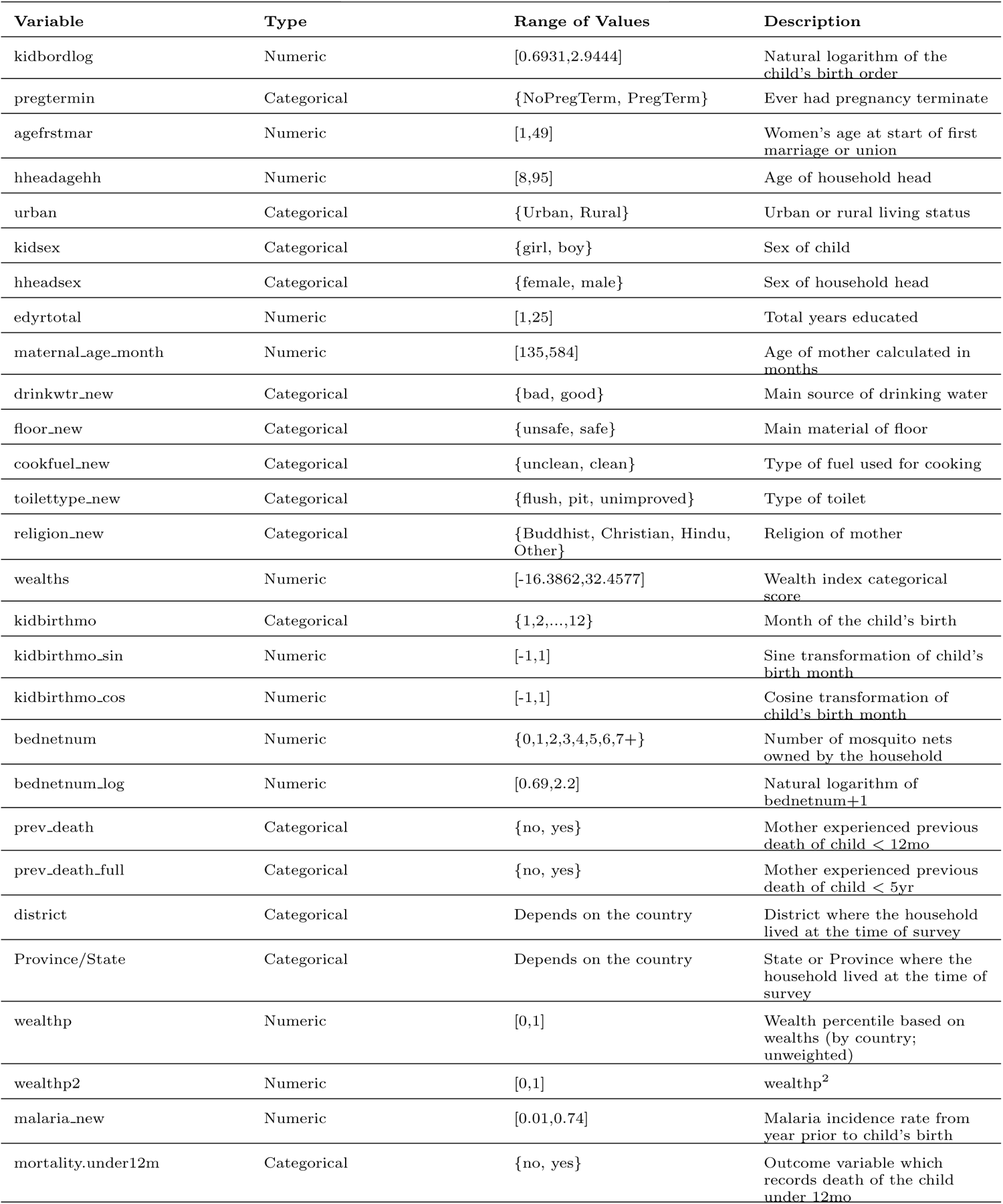
Variable information.

## 1 Country-by-Country Results

Efficiency gain is measured by the ratio of recall10 results for each algorithm compared to the recall10 results of the wealth model. Table 4 shows the average results from cross-validation on the training set, and Table 5 shows the results when the trained models are applied to the testing set.

**Table 4:**
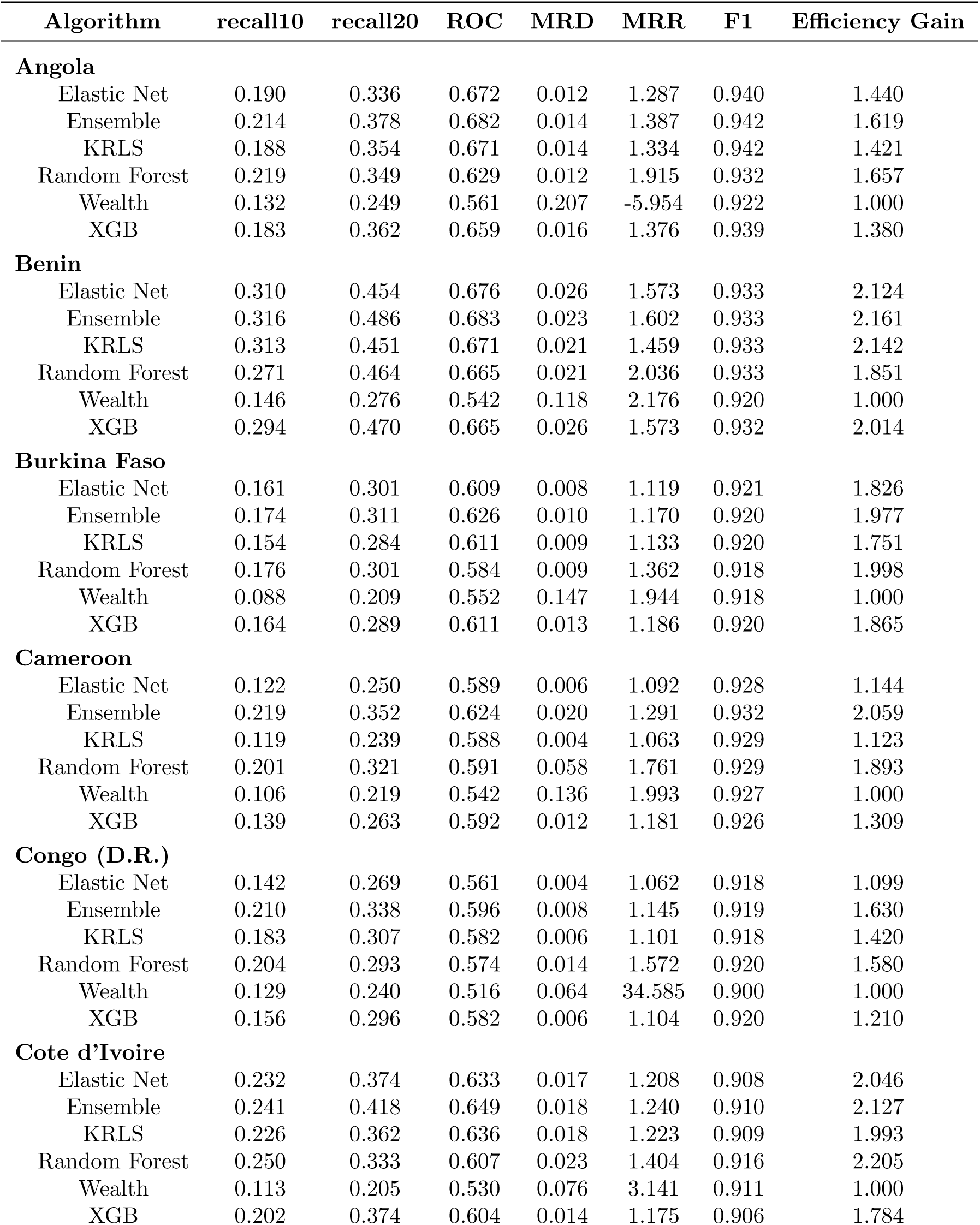
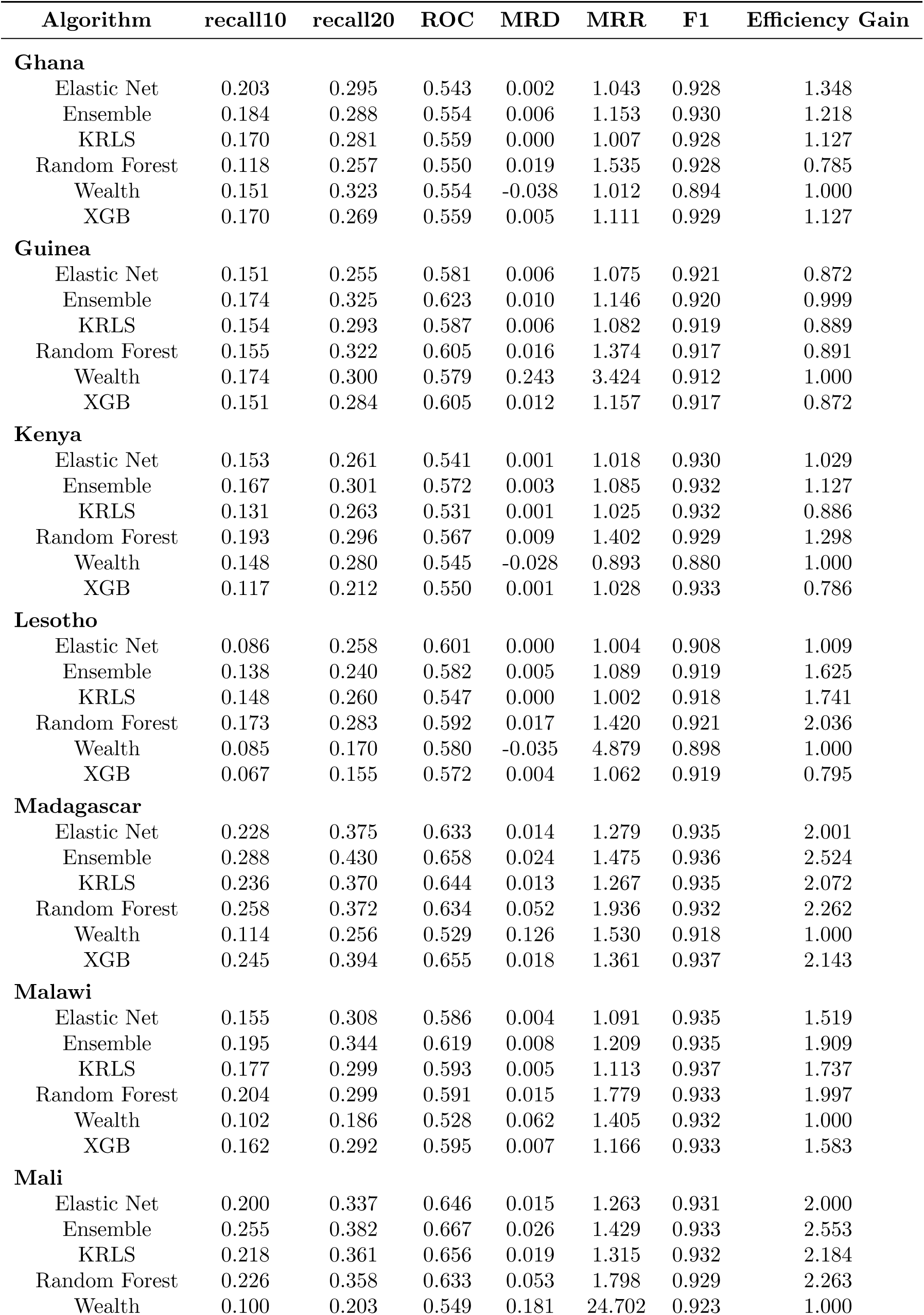
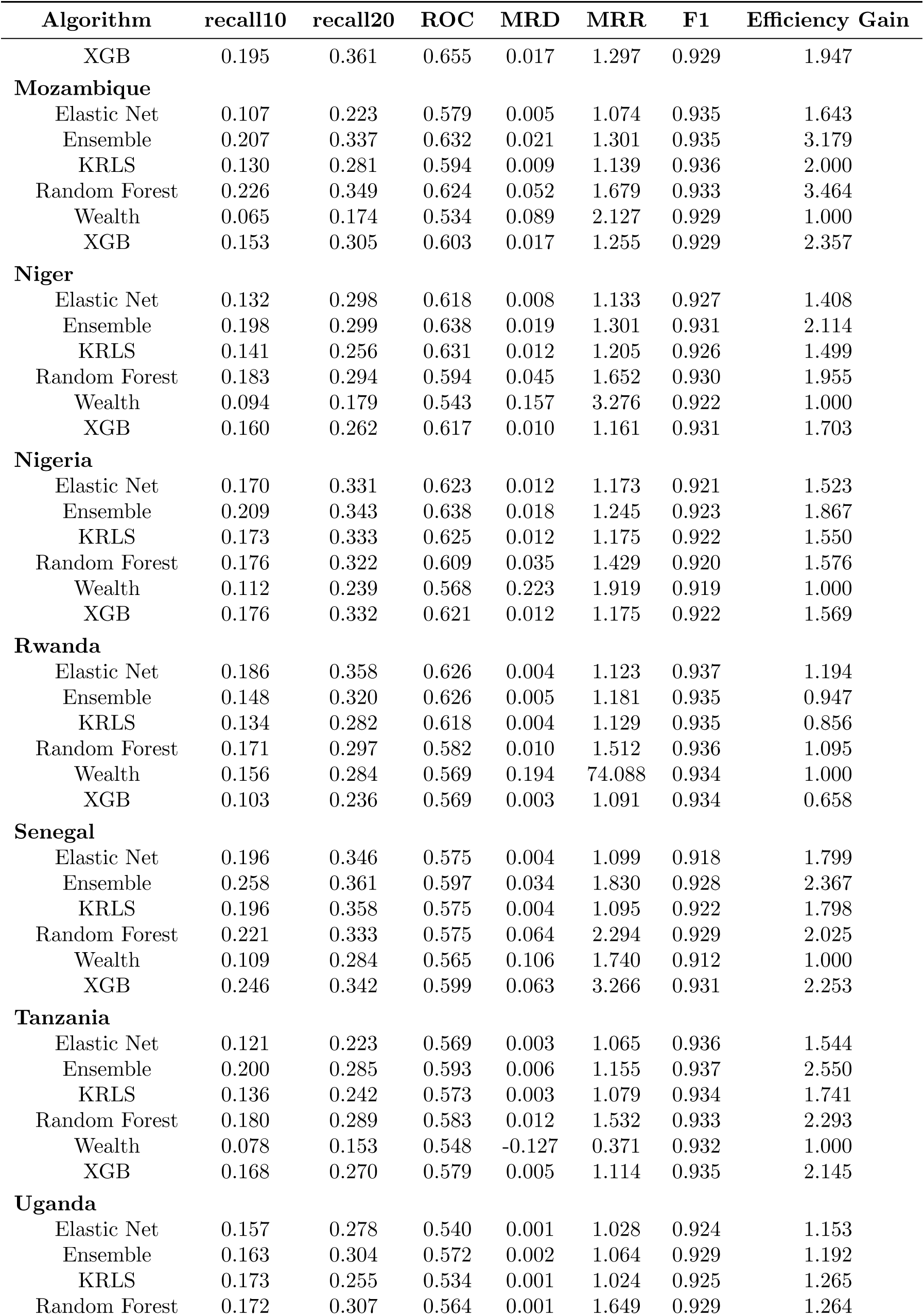
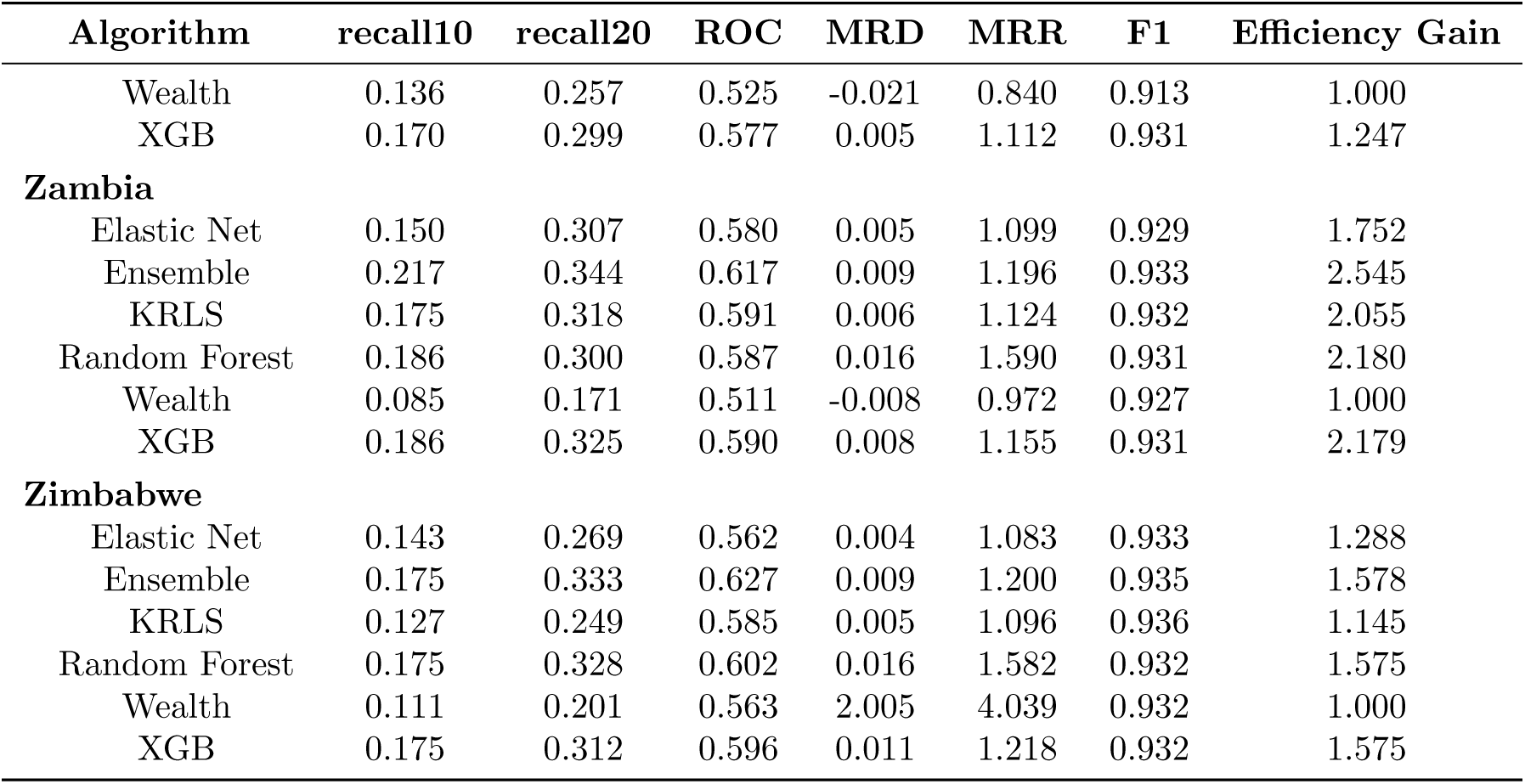
Detailed results by country, cross-validation on training set.

| Algorithm | recall10 | recall20 | ROC | MRD | MRR | F1 | Efficiency Gain |
| --- | --- | --- | --- | --- | --- | --- | --- |
| **Angola** |  |  |  |  |  |  |  |
| Elastic Net | 0.190 | 0.336 | 0.672 | 0.012 | 1.287 | 0.940 | 1.440 |
| Ensemble | 0.214 | 0.378 | 0.682 | 0.014 | 1.387 | 0.942 | 1.619 |
| KRLS | 0.188 | 0.354 | 0.671 | 0.014 | 1.334 | 0.942 | 1.421 |
| Random Forest | 0.219 | 0.349 | 0.629 | 0.012 | 1.915 | 0.932 | 1.657 |
| Wealth | 0.132 | 0.249 | 0.561 | 0.207 | -5.954 | 0.922 | 1.000 |
| XGB | 0.183 | 0.362 | 0.659 | 0.016 | 1.376 | 0.939 | 1.380 |
| **Benin** |  |  |  |  |  |  |  |
| Elastic Net | 0.310 | 0.454 | 0.676 | 0.026 | 1.573 | 0.933 | 2.124 |
| Ensemble | 0.316 | 0.486 | 0.683 | 0.023 | 1.602 | 0.933 | 2.161 |
| KRLS | 0.313 | 0.451 | 0.671 | 0.021 | 1.459 | 0.933 | 2.142 |
| Random Forest | 0.271 | 0.464 | 0.665 | 0.021 | 2.036 | 0.933 | 1.851 |
| Wealth | 0.146 | 0.276 | 0.542 | 0.118 | 2.176 | 0.920 | 1.000 |
| XGB | 0.294 | 0.470 | 0.665 | 0.026 | 1.573 | 0.932 | 2.014 |
| **Burkina Faso** |  |  |  |  |  |  |  |
| Elastic Net | 0.161 | 0.301 | 0.609 | 0.008 | 1.119 | 0.921 | 1.826 |
| Ensemble | 0.174 | 0.311 | 0.626 | 0.010 | 1.170 | 0.920 | 1.977 |
| KRLS | 0.154 | 0.284 | 0.611 | 0.009 | 1.133 | 0.920 | 1.751 |
| Random Forest | 0.176 | 0.301 | 0.584 | 0.009 | 1.362 | 0.918 | 1.998 |
| Wealth | 0.088 | 0.209 | 0.552 | 0.147 | 1.944 | 0.918 | 1.000 |
| XGB | 0.164 | 0.289 | 0.611 | 0.013 | 1.186 | 0.920 | 1.865 |
| **Cameroon** |  |  |  |  |  |  |  |
| Elastic Net | 0.122 | 0.250 | 0.589 | 0.006 | 1.092 | 0.928 | 1.144 |
| Ensemble | 0.219 | 0.352 | 0.624 | 0.020 | 1.291 | 0.932 | 2.059 |
| KRLS | 0.119 | 0.239 | 0.588 | 0.004 | 1.063 | 0.929 | 1.123 |
| Random Forest | 0.201 | 0.321 | 0.591 | 0.058 | 1.761 | 0.929 | 1.893 |
| Wealth | 0.106 | 0.219 | 0.542 | 0.136 | 1.993 | 0.927 | 1.000 |
| XGB | 0.139 | 0.263 | 0.592 | 0.012 | 1.181 | 0.926 | 1.309 |
| **Congo (D.R.)** |  |  |  |  |  |  |  |
| Elastic Net | 0.142 | 0.269 | 0.561 | 0.004 | 1.062 | 0.918 | 1.099 |
| Ensemble | 0.210 | 0.338 | 0.596 | 0.008 | 1.145 | 0.919 | 1.630 |
| KRLS | 0.183 | 0.307 | 0.582 | 0.006 | 1.101 | 0.918 | 1.420 |
| Random Forest | 0.204 | 0.293 | 0.574 | 0.014 | 1.572 | 0.920 | 1.580 |
| Wealth | 0.129 | 0.240 | 0.516 | 0.064 | 34.585 | 0.900 | 1.000 |
| XGB | 0.156 | 0.296 | 0.582 | 0.006 | 1.104 | 0.920 | 1.210 |
| **Cote d'Ivoire** |  |  |  |  |  |  |  |
| Elastic Net | 0.232 | 0.374 | 0.633 | 0.017 | 1.208 | 0.908 | 2.046 |
| Ensemble | 0.241 | 0.418 | 0.649 | 0.018 | 1.240 | 0.910 | 2.127 |
| KRLS | 0.226 | 0.362 | 0.636 | 0.018 | 1.223 | 0.909 | 1.993 |
| Random Forest | 0.250 | 0.333 | 0.607 | 0.023 | 1.404 | 0.916 | 2.205 |
| Wealth | 0.113 | 0.205 | 0.530 | 0.076 | 3.141 | 0.911 | 1.000 |
| XGB | 0.202 | 0.374 | 0.604 | 0.014 | 1.175 | 0.906 | 1.784 |
| **Ghana** |  |  |  |  |  |  |  |
| Elastic Net | 0.203 | 0.295 | 0.543 | 0.002 | 1.043 | 0.928 | 1.348 |
| Ensemble | 0.184 | 0.288 | 0.554 | 0.006 | 1.153 | 0.930 | 1.218 |
| KRLS | 0.170 | 0.281 | 0.559 | 0.000 | 1.007 | 0.928 | 1.127 |
| Random Forest | 0.118 | 0.257 | 0.550 | 0.019 | 1.535 | 0.928 | 0.785 |
| Wealth | 0.151 | 0.323 | 0.554 | -0.038 | 1.012 | 0.894 | 1.000 |
| XGB | 0.170 | 0.269 | 0.559 | 0.005 | 1.111 | 0.929 | 1.127 |
| **Guinea** |  |  |  |  |  |  |  |
| Elastic Net | 0.151 | 0.255 | 0.581 | 0.006 | 1.075 | 0.921 | 0.872 |
| Ensemble | 0.174 | 0.325 | 0.623 | 0.010 | 1.146 | 0.920 | 0.999 |
| KRLS | 0.154 | 0.293 | 0.587 | 0.006 | 1.082 | 0.919 | 0.889 |
| Random Forest | 0.155 | 0.322 | 0.605 | 0.016 | 1.374 | 0.917 | 0.891 |
| Wealth | 0.174 | 0.300 | 0.579 | 0.243 | 3.424 | 0.912 | 1.000 |
| XGB | 0.151 | 0.284 | 0.605 | 0.012 | 1.157 | 0.917 | 0.872 |
| **Kenya** |  |  |  |  |  |  |  |
| Elastic Net | 0.153 | 0.261 | 0.541 | 0.001 | 1.018 | 0.930 | 1.029 |
| Ensemble | 0.167 | 0.301 | 0.572 | 0.003 | 1.085 | 0.932 | 1.127 |
| KRLS | 0.131 | 0.263 | 0.531 | 0.001 | 1.025 | 0.932 | 0.886 |
| Random Forest | 0.193 | 0.296 | 0.567 | 0.009 | 1.402 | 0.929 | 1.298 |
| Wealth | 0.148 | 0.280 | 0.545 | -0.028 | 0.893 | 0.880 | 1.000 |
| XGB | 0.117 | 0.212 | 0.550 | 0.001 | 1.028 | 0.933 | 0.786 |
| **Lesotho** |  |  |  |  |  |  |  |
| Elastic Net | 0.086 | 0.258 | 0.601 | 0.000 | 1.004 | 0.908 | 1.009 |
| Ensemble | 0.138 | 0.240 | 0.582 | 0.005 | 1.089 | 0.919 | 1.625 |
| KRLS | 0.148 | 0.260 | 0.547 | 0.000 | 1.002 | 0.918 | 1.741 |
| Random Forest | 0.173 | 0.283 | 0.592 | 0.017 | 1.420 | 0.921 | 2.036 |
| Wealth | 0.085 | 0.170 | 0.580 | -0.035 | 4.879 | 0.898 | 1.000 |
| XGB | 0.067 | 0.155 | 0.572 | 0.004 | 1.062 | 0.919 | 0.795 |
| **Madagascar** |  |  |  |  |  |  |  |
| Elastic Net | 0.228 | 0.375 | 0.633 | 0.014 | 1.279 | 0.935 | 2.001 |
| Ensemble | 0.288 | 0.430 | 0.658 | 0.024 | 1.475 | 0.936 | 2.524 |
| KRLS | 0.236 | 0.370 | 0.644 | 0.013 | 1.267 | 0.935 | 2.072 |
| Random Forest | 0.258 | 0.372 | 0.634 | 0.052 | 1.936 | 0.932 | 2.262 |
| Wealth | 0.114 | 0.256 | 0.529 | 0.126 | 1.530 | 0.918 | 1.000 |
| XGB | 0.245 | 0.394 | 0.655 | 0.018 | 1.361 | 0.937 | 2.143 |
| **Malawi** |  |  |  |  |  |  |  |
| Elastic Net | 0.155 | 0.308 | 0.586 | 0.004 | 1.091 | 0.935 | 1.519 |
| Ensemble | 0.195 | 0.344 | 0.619 | 0.008 | 1.209 | 0.935 | 1.909 |
| KRLS | 0.177 | 0.299 | 0.593 | 0.005 | 1.113 | 0.937 | 1.737 |
| Random Forest | 0.204 | 0.299 | 0.591 | 0.015 | 1.779 | 0.933 | 1.997 |
| Wealth | 0.102 | 0.186 | 0.528 | 0.062 | 1.405 | 0.932 | 1.000 |
| XGB | 0.162 | 0.292 | 0.595 | 0.007 | 1.166 | 0.933 | 1.583 |
| **Mali** |  |  |  |  |  |  |  |
| Elastic Net | 0.200 | 0.337 | 0.646 | 0.015 | 1.263 | 0.931 | 2.000 |
| Ensemble | 0.255 | 0.382 | 0.667 | 0.026 | 1.429 | 0.933 | 2.553 |
| KRLS | 0.218 | 0.361 | 0.656 | 0.019 | 1.315 | 0.932 | 2.184 |
| Random Forest | 0.226 | 0.358 | 0.633 | 0.053 | 1.798 | 0.929 | 2.263 |
| Wealth | 0.100 | 0.203 | 0.549 | 0.181 | 24.702 | 0.923 | 1.000 |
| XGB | 0.195 | 0.361 | 0.655 | 0.017 | 1.297 | 0.929 | 1.947 |
| **Mozambique** |  |  |  |  |  |  |  |
| Elastic Net | 0.107 | 0.223 | 0.579 | 0.005 | 1.074 | 0.935 | 1.643 |
| Ensemble | 0.207 | 0.337 | 0.632 | 0.021 | 1.301 | 0.935 | 3.179 |
| KRLS | 0.130 | 0.281 | 0.594 | 0.009 | 1.139 | 0.936 | 2.000 |
| Random Forest | 0.226 | 0.349 | 0.624 | 0.052 | 1.679 | 0.933 | 3.464 |
| Wealth | 0.065 | 0.174 | 0.534 | 0.089 | 2.127 | 0.929 | 1.000 |
| XGB | 0.153 | 0.305 | 0.603 | 0.017 | 1.255 | 0.929 | 2.357 |
| **Niger** |  |  |  |  |  |  |  |
| Elastic Net | 0.132 | 0.298 | 0.618 | 0.008 | 1.133 | 0.927 | 1.408 |
| Ensemble | 0.198 | 0.299 | 0.638 | 0.019 | 1.301 | 0.931 | 2.114 |
| KRLS | 0.141 | 0.256 | 0.631 | 0.012 | 1.205 | 0.926 | 1.499 |
| Random Forest | 0.183 | 0.294 | 0.594 | 0.045 | 1.652 | 0.930 | 1.955 |
| Wealth | 0.094 | 0.179 | 0.543 | 0.157 | 3.276 | 0.922 | 1.000 |
| XGB | 0.160 | 0.262 | 0.617 | 0.010 | 1.161 | 0.931 | 1.703 |
| **Nigeria** |  |  |  |  |  |  |  |
| Elastic Net | 0.170 | 0.331 | 0.623 | 0.012 | 1.173 | 0.921 | 1.523 |
| Ensemble | 0.209 | 0.343 | 0.638 | 0.018 | 1.245 | 0.923 | 1.867 |
| KRLS | 0.173 | 0.333 | 0.625 | 0.012 | 1.175 | 0.922 | 1.550 |
| Random Forest | 0.176 | 0.322 | 0.609 | 0.035 | 1.429 | 0.920 | 1.576 |
| Wealth | 0.112 | 0.239 | 0.568 | 0.223 | 1.919 | 0.919 | 1.000 |
| XGB | 0.176 | 0.332 | 0.621 | 0.012 | 1.175 | 0.922 | 1.569 |
| **Rwanda** |  |  |  |  |  |  |  |
| Elastic Net | 0.186 | 0.358 | 0.626 | 0.004 | 1.123 | 0.937 | 1.194 |
| Ensemble | 0.148 | 0.320 | 0.626 | 0.005 | 1.181 | 0.935 | 0.947 |
| KRLS | 0.134 | 0.282 | 0.618 | 0.004 | 1.129 | 0.935 | 0.856 |
| Random Forest | 0.171 | 0.297 | 0.582 | 0.010 | 1.512 | 0.936 | 1.095 |
| Wealth | 0.156 | 0.284 | 0.569 | 0.194 | 74.088 | 0.934 | 1.000 |
| XGB | 0.103 | 0.236 | 0.569 | 0.003 | 1.091 | 0.934 | 0.658 |
| **Senegal** |  |  |  |  |  |  |  |
| Elastic Net | 0.196 | 0.346 | 0.575 | 0.004 | 1.099 | 0.918 | 1.799 |
| Ensemble | 0.258 | 0.361 | 0.597 | 0.034 | 1.830 | 0.928 | 2.367 |
| KRLS | 0.196 | 0.358 | 0.575 | 0.004 | 1.095 | 0.922 | 1.798 |
| Random Forest | 0.221 | 0.333 | 0.575 | 0.064 | 2.294 | 0.929 | 2.025 |
| Wealth | 0.109 | 0.284 | 0.565 | 0.106 | 1.740 | 0.912 | 1.000 |
| XGB | 0.246 | 0.342 | 0.599 | 0.063 | 3.266 | 0.931 | 2.253 |
| **Tanzania** |  |  |  |  |  |  |  |
| Elastic Net | 0.121 | 0.223 | 0.569 | 0.003 | 1.065 | 0.936 | 1.544 |
| Ensemble | 0.200 | 0.285 | 0.593 | 0.006 | 1.155 | 0.937 | 2.550 |
| KRLS | 0.136 | 0.242 | 0.573 | 0.003 | 1.079 | 0.934 | 1.741 |
| Random Forest | 0.180 | 0.289 | 0.583 | 0.012 | 1.532 | 0.933 | 2.293 |
| Wealth | 0.078 | 0.153 | 0.548 | -0.127 | 0.371 | 0.932 | 1.000 |
| XGB | 0.168 | 0.270 | 0.579 | 0.005 | 1.114 | 0.935 | 2.145 |
| **Uganda** |  |  |  |  |  |  |  |
| Elastic Net | 0.157 | 0.278 | 0.540 | 0.001 | 1.028 | 0.924 | 1.153 |
| Ensemble | 0.163 | 0.304 | 0.572 | 0.002 | 1.064 | 0.929 | 1.192 |
| KRLS | 0.173 | 0.255 | 0.534 | 0.001 | 1.024 | 0.925 | 1.265 |
| Random Forest | 0.172 | 0.307 | 0.564 | 0.001 | 1.649 | 0.929 | 1.264 |
| Wealth | 0.136 | 0.257 | 0.525 | -0.021 | 0.840 | 0.913 | 1.000 |
| XGB | 0.170 | 0.299 | 0.577 | 0.005 | 1.112 | 0.931 | 1.247 |
| **Zambia** |  |  |  |  |  |  |  |
| Elastic Net | 0.150 | 0.307 | 0.580 | 0.005 | 1.099 | 0.929 | 1.752 |
| Ensemble | 0.217 | 0.344 | 0.617 | 0.009 | 1.196 | 0.933 | 2.545 |
| KRLS | 0.175 | 0.318 | 0.591 | 0.006 | 1.124 | 0.932 | 2.055 |
| Random Forest | 0.186 | 0.300 | 0.587 | 0.016 | 1.590 | 0.931 | 2.180 |
| Wealth | 0.085 | 0.171 | 0.511 | -0.008 | 0.972 | 0.927 | 1.000 |
| XGB | 0.186 | 0.325 | 0.590 | 0.008 | 1.155 | 0.931 | 2.179 |
| **Zimbabwe** |  |  |  |  |  |  |  |
| Elastic Net | 0.143 | 0.269 | 0.562 | 0.004 | 1.083 | 0.933 | 1.288 |
| Ensemble | 0.175 | 0.333 | 0.627 | 0.009 | 1.200 | 0.935 | 1.578 |
| KRLS | 0.127 | 0.249 | 0.585 | 0.005 | 1.096 | 0.936 | 1.145 |
| Random Forest | 0.175 | 0.328 | 0.602 | 0.016 | 1.582 | 0.932 | 1.575 |
| Wealth | 0.111 | 0.201 | 0.563 | 2.005 | 4.039 | 0.932 | 1.000 |
| XGB | 0.175 | 0.312 | 0.596 | 0.011 | 1.218 | 0.932 | 1.575 |

**Table 5:**
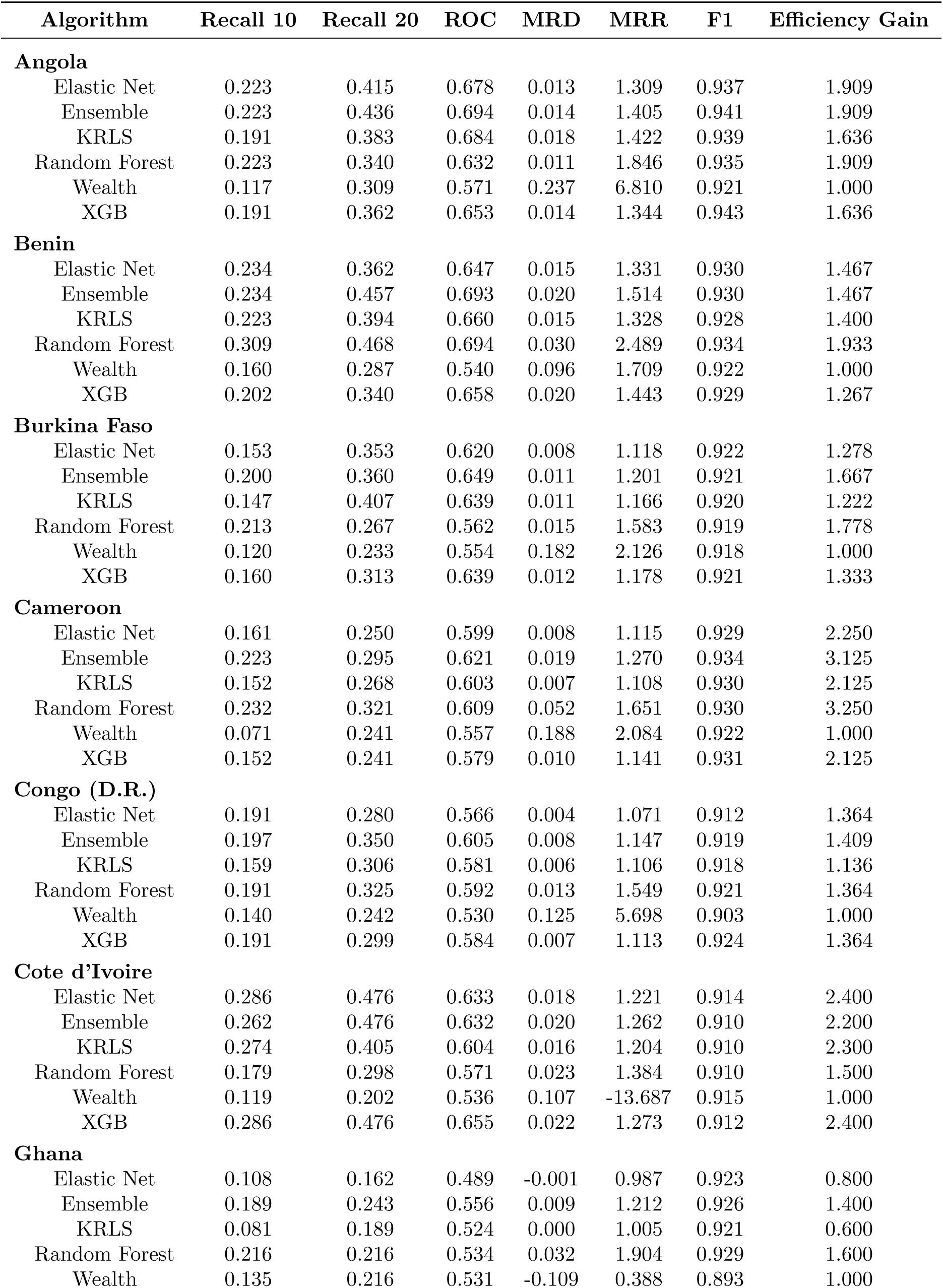
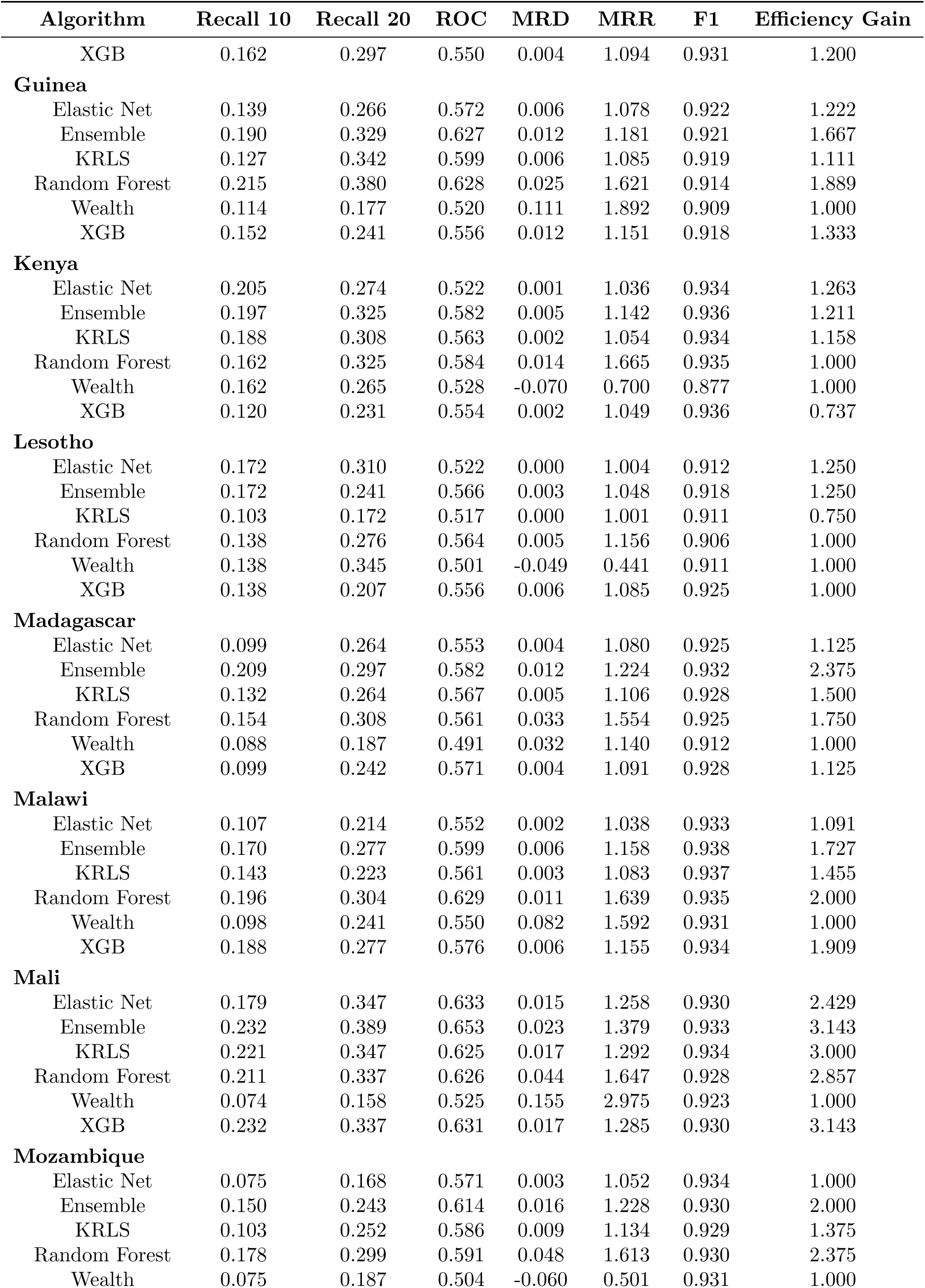
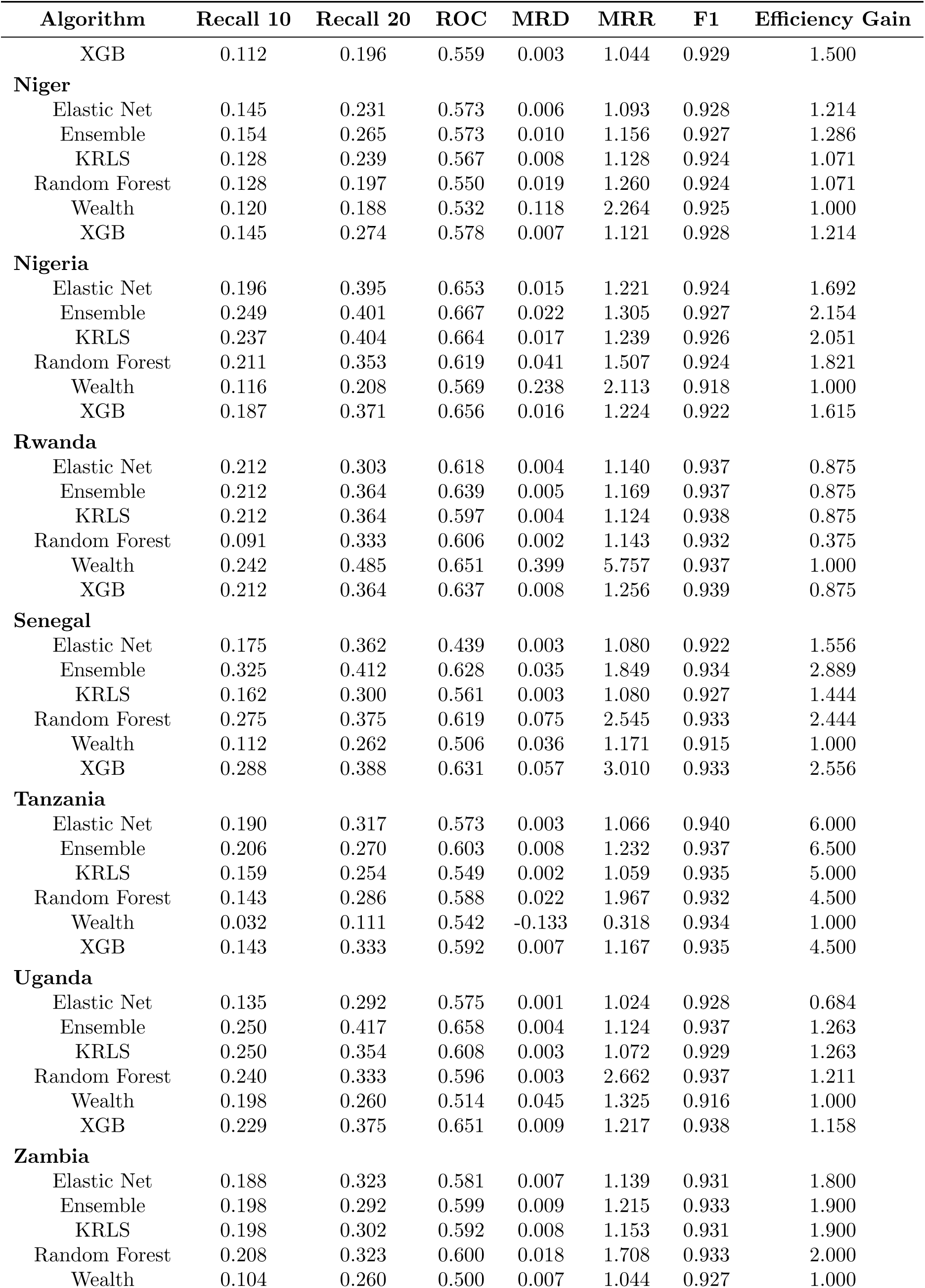
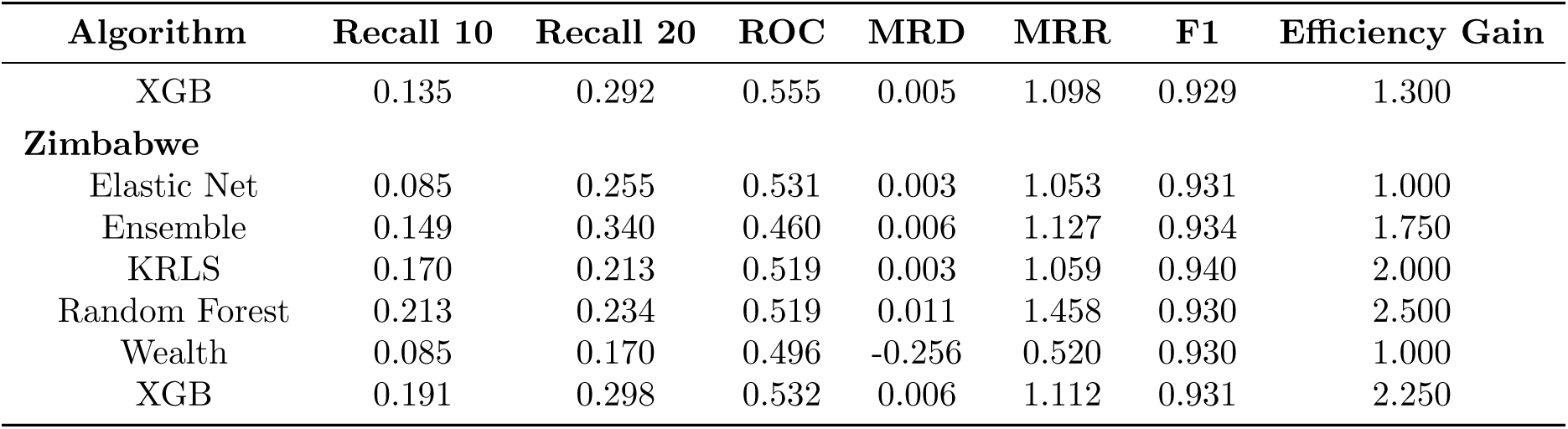
Detailed results by country, test set.

| Algorithm | Recall 10 | Recall 20 | ROC | MRD | MRR | F1 | Efficiency Gain |
| --- | --- | --- | --- | --- | --- | --- | --- |
| **Angola** |  |  |  |  |  |  |  |
| Elastic Net | 0.223 | 0.415 | 0.678 | 0.013 | 1.309 | 0.937 | 1.909 |
| Ensemble | 0.223 | 0.436 | 0.694 | 0.014 | 1.405 | 0.941 | 1.909 |
| KRLS | 0.191 | 0.383 | 0.684 | 0.018 | 1.422 | 0.939 | 1.636 |
| Random Forest | 0.223 | 0.340 | 0.632 | 0.011 | 1.846 | 0.935 | 1.909 |
| Wealth | 0.117 | 0.309 | 0.571 | 0.237 | 6.810 | 0.921 | 1.000 |
| XGB | 0.191 | 0.362 | 0.653 | 0.014 | 1.344 | 0.943 | 1.636 |
| **Benin** |  |  |  |  |  |  |  |
| Elastic Net | 0.234 | 0.362 | 0.647 | 0.015 | 1.331 | 0.930 | 1.467 |
| Ensemble | 0.234 | 0.457 | 0.693 | 0.020 | 1.514 | 0.930 | 1.467 |
| KRLS | 0.223 | 0.394 | 0.660 | 0.015 | 1.328 | 0.928 | 1.400 |
| Random Forest | 0.309 | 0.468 | 0.694 | 0.030 | 2.489 | 0.934 | 1.933 |
| Wealth | 0.160 | 0.287 | 0.540 | 0.096 | 1.709 | 0.922 | 1.000 |
| XGB | 0.202 | 0.340 | 0.658 | 0.020 | 1.443 | 0.929 | 1.267 |
| **Burkina Faso** |  |  |  |  |  |  |  |
| Elastic Net | 0.153 | 0.353 | 0.620 | 0.008 | 1.118 | 0.922 | 1.278 |
| Ensemble | 0.200 | 0.360 | 0.649 | 0.011 | 1.201 | 0.921 | 1.667 |
| KRLS | 0.147 | 0.407 | 0.639 | 0.011 | 1.166 | 0.920 | 1.222 |
| Random Forest | 0.213 | 0.267 | 0.562 | 0.015 | 1.583 | 0.919 | 1.778 |
| Wealth | 0.120 | 0.233 | 0.554 | 0.182 | 2.126 | 0.918 | 1.000 |
| XGB | 0.160 | 0.313 | 0.639 | 0.012 | 1.178 | 0.921 | 1.333 |
| **Cameroon** |  |  |  |  |  |  |  |
| Elastic Net | 0.161 | 0.250 | 0.599 | 0.008 | 1.115 | 0.929 | 2.250 |
| Ensemble | 0.223 | 0.295 | 0.621 | 0.019 | 1.270 | 0.934 | 3.125 |
| KRLS | 0.152 | 0.268 | 0.603 | 0.007 | 1.108 | 0.930 | 2.125 |
| Random Forest | 0.232 | 0.321 | 0.609 | 0.052 | 1.651 | 0.930 | 3.250 |
| Wealth | 0.071 | 0.241 | 0.557 | 0.188 | 2.084 | 0.922 | 1.000 |
| XGB | 0.152 | 0.241 | 0.579 | 0.010 | 1.141 | 0.931 | 2.125 |
| **Congo (D.R.)** |  |  |  |  |  |  |  |
| Elastic Net | 0.191 | 0.280 | 0.566 | 0.004 | 1.071 | 0.912 | 1.364 |
| Ensemble | 0.197 | 0.350 | 0.605 | 0.008 | 1.147 | 0.919 | 1.409 |
| KRLS | 0.159 | 0.306 | 0.581 | 0.006 | 1.106 | 0.918 | 1.136 |
| Random Forest | 0.191 | 0.325 | 0.592 | 0.013 | 1.549 | 0.921 | 1.364 |
| Wealth | 0.140 | 0.242 | 0.530 | 0.125 | 5.698 | 0.903 | 1.000 |
| XGB | 0.191 | 0.299 | 0.584 | 0.007 | 1.113 | 0.924 | 1.364 |
| **Cote d'Ivoire** |  |  |  |  |  |  |  |
| Elastic Net | 0.286 | 0.476 | 0.633 | 0.018 | 1.221 | 0.914 | 2.400 |
| Ensemble | 0.262 | 0.476 | 0.632 | 0.020 | 1.262 | 0.910 | 2.200 |
| KRLS | 0.274 | 0.405 | 0.604 | 0.016 | 1.204 | 0.910 | 2.300 |
| Random Forest | 0.179 | 0.298 | 0.571 | 0.023 | 1.384 | 0.910 | 1.500 |
| Wealth | 0.119 | 0.202 | 0.536 | 0.107 | -13.687 | 0.915 | 1.000 |
| XGB | 0.286 | 0.476 | 0.655 | 0.022 | 1.273 | 0.912 | 2.400 |
| **Ghana** |  |  |  |  |  |  |  |
| Elastic Net | 0.108 | 0.162 | 0.489 | -0.001 | 0.987 | 0.923 | 0.800 |
| Ensemble | 0.189 | 0.243 | 0.556 | 0.009 | 1.212 | 0.926 | 1.400 |
| KRLS | 0.081 | 0.189 | 0.524 | 0.000 | 1.005 | 0.921 | 0.600 |
| Random Forest | 0.216 | 0.216 | 0.534 | 0.032 | 1.904 | 0.929 | 1.600 |
| Wealth | 0.135 | 0.216 | 0.531 | -0.109 | 0.388 | 0.893 | 1.000 |
| XGB | 0.162 | 0.297 | 0.550 | 0.004 | 1.094 | 0.931 | 1.200 |
| **Guinea** |  |  |  |  |  |  |  |
| Elastic Net | 0.139 | 0.266 | 0.572 | 0.006 | 1.078 | 0.922 | 1.222 |
| Ensemble | 0.190 | 0.329 | 0.627 | 0.012 | 1.181 | 0.921 | 1.667 |
| KRLS | 0.127 | 0.342 | 0.599 | 0.006 | 1.085 | 0.919 | 1.111 |
| Random Forest | 0.215 | 0.380 | 0.628 | 0.025 | 1.621 | 0.914 | 1.889 |
| Wealth | 0.114 | 0.177 | 0.520 | 0.111 | 1.892 | 0.909 | 1.000 |
| XGB | 0.152 | 0.241 | 0.556 | 0.012 | 1.151 | 0.918 | 1.333 |
| **Kenya** |  |  |  |  |  |  |  |
| Elastic Net | 0.205 | 0.274 | 0.522 | 0.001 | 1.036 | 0.934 | 1.263 |
| Ensemble | 0.197 | 0.325 | 0.582 | 0.005 | 1.142 | 0.936 | 1.211 |
| KRLS | 0.188 | 0.308 | 0.563 | 0.002 | 1.054 | 0.934 | 1.158 |
| Random Forest | 0.162 | 0.325 | 0.584 | 0.014 | 1.665 | 0.935 | 1.000 |
| Wealth | 0.162 | 0.265 | 0.528 | -0.070 | 0.700 | 0.877 | 1.000 |
| XGB | 0.120 | 0.231 | 0.554 | 0.002 | 1.049 | 0.936 | 0.737 |
| **Lesotho** |  |  |  |  |  |  |  |
| Elastic Net | 0.172 | 0.310 | 0.522 | 0.000 | 1.004 | 0.912 | 1.250 |
| Ensemble | 0.172 | 0.241 | 0.566 | 0.003 | 1.048 | 0.918 | 1.250 |
| KRLS | 0.103 | 0.172 | 0.517 | 0.000 | 1.001 | 0.911 | 0.750 |
| Random Forest | 0.138 | 0.276 | 0.564 | 0.005 | 1.156 | 0.906 | 1.000 |
| Wealth | 0.138 | 0.345 | 0.501 | -0.049 | 0.441 | 0.911 | 1.000 |
| XGB | 0.138 | 0.207 | 0.556 | 0.006 | 1.085 | 0.925 | 1.000 |
| **Madagascar** |  |  |  |  |  |  |  |
| Elastic Net | 0.099 | 0.264 | 0.553 | 0.004 | 1.080 | 0.925 | 1.125 |
| Ensemble | 0.209 | 0.297 | 0.582 | 0.012 | 1.224 | 0.932 | 2.375 |
| KRLS | 0.132 | 0.264 | 0.567 | 0.005 | 1.106 | 0.928 | 1.500 |
| Random Forest | 0.154 | 0.308 | 0.561 | 0.033 | 1.554 | 0.925 | 1.750 |
| Wealth | 0.088 | 0.187 | 0.491 | 0.032 | 1.140 | 0.912 | 1.000 |
| XGB | 0.099 | 0.242 | 0.571 | 0.004 | 1.091 | 0.928 | 1.125 |
| **Malawi** |  |  |  |  |  |  |  |
| Elastic Net | 0.107 | 0.214 | 0.552 | 0.002 | 1.038 | 0.933 | 1.091 |
| Ensemble | 0.170 | 0.277 | 0.599 | 0.006 | 1.158 | 0.938 | 1.727 |
| KRLS | 0.143 | 0.223 | 0.561 | 0.003 | 1.083 | 0.937 | 1.455 |
| Random Forest | 0.196 | 0.304 | 0.629 | 0.011 | 1.639 | 0.935 | 2.000 |
| Wealth | 0.098 | 0.241 | 0.550 | 0.082 | 1.592 | 0.931 | 1.000 |
| XGB | 0.188 | 0.277 | 0.576 | 0.006 | 1.155 | 0.934 | 1.909 |
| **Mali** |  |  |  |  |  |  |  |
| Elastic Net | 0.179 | 0.347 | 0.633 | 0.015 | 1.258 | 0.930 | 2.429 |
| Ensemble | 0.232 | 0.389 | 0.653 | 0.023 | 1.379 | 0.933 | 3.143 |
| KRLS | 0.221 | 0.347 | 0.625 | 0.017 | 1.292 | 0.934 | 3.000 |
| Random Forest | 0.211 | 0.337 | 0.626 | 0.044 | 1.647 | 0.928 | 2.857 |
| Wealth | 0.074 | 0.158 | 0.525 | 0.155 | 2.975 | 0.923 | 1.000 |
| XGB | 0.232 | 0.337 | 0.631 | 0.017 | 1.285 | 0.930 | 3.143 |
| **Mozambique** |  |  |  |  |  |  |  |
| Elastic Net | 0.075 | 0.168 | 0.571 | 0.003 | 1.052 | 0.934 | 1.000 |
| Ensemble | 0.150 | 0.243 | 0.614 | 0.016 | 1.228 | 0.930 | 2.000 |
| KRLS | 0.103 | 0.252 | 0.586 | 0.009 | 1.134 | 0.929 | 1.375 |
| Random Forest | 0.178 | 0.299 | 0.591 | 0.048 | 1.613 | 0.930 | 2.375 |
| Wealth | 0.075 | 0.187 | 0.504 | -0.060 | 0.501 | 0.931 | 1.000 |
| XGB | 0.112 | 0.196 | 0.559 | 0.003 | 1.044 | 0.929 | 1.500 |
| **Niger** |  |  |  |  |  |  |  |
| Elastic Net | 0.145 | 0.231 | 0.573 | 0.006 | 1.093 | 0.928 | 1.214 |
| Ensemble | 0.154 | 0.265 | 0.573 | 0.010 | 1.156 | 0.927 | 1.286 |
| KRLS | 0.128 | 0.239 | 0.567 | 0.008 | 1.128 | 0.924 | 1.071 |
| Random Forest | 0.128 | 0.197 | 0.550 | 0.019 | 1.260 | 0.924 | 1.071 |
| Wealth | 0.120 | 0.188 | 0.532 | 0.118 | 2.264 | 0.925 | 1.000 |
| XGB | 0.145 | 0.274 | 0.578 | 0.007 | 1.121 | 0.928 | 1.214 |
| **Nigeria** |  |  |  |  |  |  |  |
| Elastic Net | 0.196 | 0.395 | 0.653 | 0.015 | 1.221 | 0.924 | 1.692 |
| Ensemble | 0.249 | 0.401 | 0.667 | 0.022 | 1.305 | 0.927 | 2.154 |
| KRLS | 0.237 | 0.404 | 0.664 | 0.017 | 1.239 | 0.926 | 2.051 |
| Random Forest | 0.211 | 0.353 | 0.619 | 0.041 | 1.507 | 0.924 | 1.821 |
| Wealth | 0.116 | 0.208 | 0.569 | 0.238 | 2.113 | 0.918 | 1.000 |
| XGB | 0.187 | 0.371 | 0.656 | 0.016 | 1.224 | 0.922 | 1.615 |
| **Rwanda** |  |  |  |  |  |  |  |
| Elastic Net | 0.212 | 0.303 | 0.618 | 0.004 | 1.140 | 0.937 | 0.875 |
| Ensemble | 0.212 | 0.364 | 0.639 | 0.005 | 1.169 | 0.937 | 0.875 |
| KRLS | 0.212 | 0.364 | 0.597 | 0.004 | 1.124 | 0.938 | 0.875 |
| Random Forest | 0.091 | 0.333 | 0.606 | 0.002 | 1.143 | 0.932 | 0.375 |
| Wealth | 0.242 | 0.485 | 0.651 | 0.399 | 5.757 | 0.937 | 1.000 |
| XGB | 0.212 | 0.364 | 0.637 | 0.008 | 1.256 | 0.939 | 0.875 |
| **Senegal** |  |  |  |  |  |  |  |
| Elastic Net | 0.175 | 0.362 | 0.439 | 0.003 | 1.080 | 0.922 | 1.556 |
| Ensemble | 0.325 | 0.412 | 0.628 | 0.035 | 1.849 | 0.934 | 2.889 |
| KRLS | 0.162 | 0.300 | 0.561 | 0.003 | 1.080 | 0.927 | 1.444 |
| Random Forest | 0.275 | 0.375 | 0.619 | 0.075 | 2.545 | 0.933 | 2.444 |
| Wealth | 0.112 | 0.262 | 0.506 | 0.036 | 1.171 | 0.915 | 1.000 |
| XGB | 0.288 | 0.388 | 0.631 | 0.057 | 3.010 | 0.933 | 2.556 |
| **Tanzania** |  |  |  |  |  |  |  |
| Elastic Net | 0.190 | 0.317 | 0.573 | 0.003 | 1.066 | 0.940 | 6.000 |
| Ensemble | 0.206 | 0.270 | 0.603 | 0.008 | 1.232 | 0.937 | 6.500 |
| KRLS | 0.159 | 0.254 | 0.549 | 0.002 | 1.059 | 0.935 | 5.000 |
| Random Forest | 0.143 | 0.286 | 0.588 | 0.022 | 1.967 | 0.932 | 4.500 |
| Wealth | 0.032 | 0.111 | 0.542 | -0.133 | 0.318 | 0.934 | 1.000 |
| XGB | 0.143 | 0.333 | 0.592 | 0.007 | 1.167 | 0.935 | 4.500 |
| **Uganda** |  |  |  |  |  |  |  |
| Elastic Net | 0.135 | 0.292 | 0.575 | 0.001 | 1.024 | 0.928 | 0.684 |
| Ensemble | 0.250 | 0.417 | 0.658 | 0.004 | 1.124 | 0.937 | 1.263 |
| KRLS | 0.250 | 0.354 | 0.608 | 0.003 | 1.072 | 0.929 | 1.263 |
| Random Forest | 0.240 | 0.333 | 0.596 | 0.003 | 2.662 | 0.937 | 1.211 |
| Wealth | 0.198 | 0.260 | 0.514 | 0.045 | 1.325 | 0.916 | 1.000 |
| XGB | 0.229 | 0.375 | 0.651 | 0.009 | 1.217 | 0.938 | 1.158 |
| **Zambia** |  |  |  |  |  |  |  |
| Elastic Net | 0.188 | 0.323 | 0.581 | 0.007 | 1.139 | 0.931 | 1.800 |
| Ensemble | 0.198 | 0.292 | 0.599 | 0.009 | 1.215 | 0.933 | 1.900 |
| KRLS | 0.198 | 0.302 | 0.592 | 0.008 | 1.153 | 0.931 | 1.900 |
| Random Forest | 0.208 | 0.323 | 0.600 | 0.018 | 1.708 | 0.933 | 2.000 |
| Wealth | 0.104 | 0.260 | 0.500 | 0.007 | 1.044 | 0.927 | 1.000 |
| XGB | 0.135 | 0.292 | 0.555 | 0.005 | 1.098 | 0.929 | 1.300 |
| **Zimbabwe** |  |  |  |  |  |  |  |
| Elastic Net | 0.085 | 0.255 | 0.531 | 0.003 | 1.053 | 0.931 | 1.000 |
| Ensemble | 0.149 | 0.340 | 0.460 | 0.006 | 1.127 | 0.934 | 1.750 |
| KRLS | 0.170 | 0.213 | 0.519 | 0.003 | 1.059 | 0.940 | 2.000 |
| Random Forest | 0.213 | 0.234 | 0.519 | 0.011 | 1.458 | 0.930 | 2.500 |
| Wealth | 0.085 | 0.170 | 0.496 | -0.256 | 0.520 | 0.930 | 1.000 |
| XGB | 0.191 | 0.298 | 0.532 | 0.006 | 1.112 | 0.931 | 2.250 |

## Footnotes

1 We thank an anonymous reviewer for noting that such an analysis neglects possible spillovers, such as when saving one child’s life could increase a nearby child’s chance of survival. Such spillovers may be reasonably likely, for example, in the case of infectious diseases. Computing the net effect of an intervention under such spillover would require additional structure and assumptions on the problem. It is noteworthy, however, that because early mortality is statistically rare, the effect of such spillover effects on the net lives saved can only be non-negligible where that spillover occurs through clusters of births that share very high-risk levels. This is possible if, indeed risk levels might be highly concentrated (e.g., in particular villages). In this case, effective interventions well targeted to such high risk areas would enjoy an even greater life-saving benefit due to this spillover.

## Notes

### Competing Interest Statement

The authors have declared no competing interest.

### Funding Statement

CCPR Population Research Infrastructure Grant P2C from NICHD P2C-HD041022

### Author Declarations

This paper does not involve human subjects

### Summary of Updates

No significant changes.

